# Using brain cell-type-specific protein interactomes to interpret genetic data in schizophrenia

**DOI:** 10.1101/2021.10.07.21264568

**Authors:** Yu-Han H. Hsu, Eugeniu Nacu, Ruize Liu, Greta Pintacuda, April Kim, Kalliopi Tsafou, Natalie Petrossian, William Crotty, Jung Min Suh, Jackson Riseman, Jacqueline M. Martin, Julia C. Biagini, Joshua K.T. Ching, Edyta Malolepsza, Taibo Li, Tarjinder Singh, Tian Ge, Shawn B. Egri, Benjamin Tanenbaum, Caroline R. Stanclift, Annie M. Apffel, Schizophrenia Working Group of the Psychiatric Genomics Consortium, Stanley Global Asia Initiatives, Steven A. Carr, Monica Schenone, Jake Jaffe, Nadine Fornelos, Hailiang Huang, Kevin C. Eggan, Kasper Lage

## Abstract

Genetics have nominated many schizophrenia risk genes that lack functional interpretation. To empower such interpretation, we executed interaction proteomics for six risk genes in human induced neurons and found the resulting protein network to be enriched for common variant risk of schizophrenia in Europeans and East Asians. The network is down-regulated in layer 5/6 cortical neurons of patients and can complement fine-mapping and eQTL data to prioritize additional genes in GWAS loci. A sub-network centered on HCN1 is enriched for common variant risk and also contains proteins (HCN4 and AKAP11) enriched for rare protein-truncating mutations in patients with schizophrenia and bipolar disease. Our findings establish brain cell-type-specific interactomes as an organizing framework to facilitate interpretation of genetic and transcriptomic data in schizophrenia and psychiatric diseases.

**One Sentence Summary:** Neuronal protein interactomes is an organizing framework for integrating genetic and transcriptomic data in schizophrenia.

## Main Text

Schizophrenia is a debilitating psychiatric disorder with a strong genetic component occurring in ∼0.3% of the global population with severe repercussions for patients, families, and society (*1, 2*). The last years have seen large advances in mapping the genetic architecture of disease and in generating catalogs of risk variants and genes. These efforts have revealed a highly polygenic disorder with contributions from hundreds of genes and rare and common variants across a wide spectrum of allele frequencies (*3–7*). Genetic risk of schizophrenia correlates between diverse populations such as Europeans and East Asians (*8*), suggesting that a discrete and shared set of cellular networks and pathways at the protein level (we will collectively refer to these as networks, hereafter) in cell types of the human brain are involved in its biology. Developing paradigms for systematically using genetic information as an entry point to investigate these brain cell-type-specific networks is therefore a key goal of psychiatry and genetics. Here, we aimed to bring together the newest developments in genetics, single-cell RNA sequencing, neuronal cell modeling, and proteomics to better understand schizophrenia at the level of brain cell-type-specific protein interactomes.

Analyses of postmortem brains from patients with schizophrenia (*9*) and integration of genetic information with RNA sequencing data from human and mouse brains (*10–12*) have converged on cortical excitatory neurons as a key biological conduit of genetically encoded risk. While other brain cell types may also be relevant to schizophrenia, cortical excitatory neurons have emerged as a pivotal cell type in which to initiate systematic mapping of cellular protein networks implicated by genetics and transcriptomics in schizophrenia. A recent study showed that adding extrinsic neuronal patterning to pluripotent stem cells (PSCs) overexpressing *NGN2* generates glutamatergic patterned induced neurons (iNs) that behave like cortical excitatory neurons at the molecular, morphological, and functional levels (*13, 14*). By adapting this protocol, we were able to develop a scalable workflow to routinely produce billions of iNs in homogeneous cell populations. Combined with the availability of robust and reproducible genetic datasets in schizophrenia, this creates a unique opportunity to systematically investigate intracellular protein interaction networks of schizophrenia risk genes in a relevant human cellular context. While other sources of neurons (e.g., murine brains, human postmortem brains, spheroids, or organoids) should be considered in the future, we started with iNs because this design choice minimizes experimental noise and biochemical artifacts introduced by a mixture of different cell types and produces a dataset that is maximally biologically and genetically interpretable.

To identify schizophrenia risk genes as the basis of our interactome experiments we designed and executed a three-step procedure (**Fig. 1A** and **Data S1**). First, we identified 445 genes in previously reported genome-wide significant loci from the Psychiatric Genomics Consortium (PGC) genome-wide association study (GWAS; phase 2) of 37K schizophrenia cases and 113K controls (*3*) (Set 1). Second, we filtered this set to 37 genes within single protein-coding gene loci (Set 2). Third, we integrated data from orthogonal genetic studies (e.g., high-density genotyping, exome sequencing, and earlier targeted studies of individual genes; **Materials and Methods** and **Data S1**) to identify a subset of 10 genes supported by multiple independent lines of evidence (Set 3): *CACNA1C*, *CACNB2*, *CSMD1*, *CUL3*, *GRIN2A*, *HCN1*, *RIMS1*, *SATB2, TCF4*, and *ZNF804A*. We additionally included *SYNGAP1* in the major histocompatibility complex (MHC) region in all three sets due to strong orthogonal evidence for its involvement in schizophrenia.

**Fig. 1.**
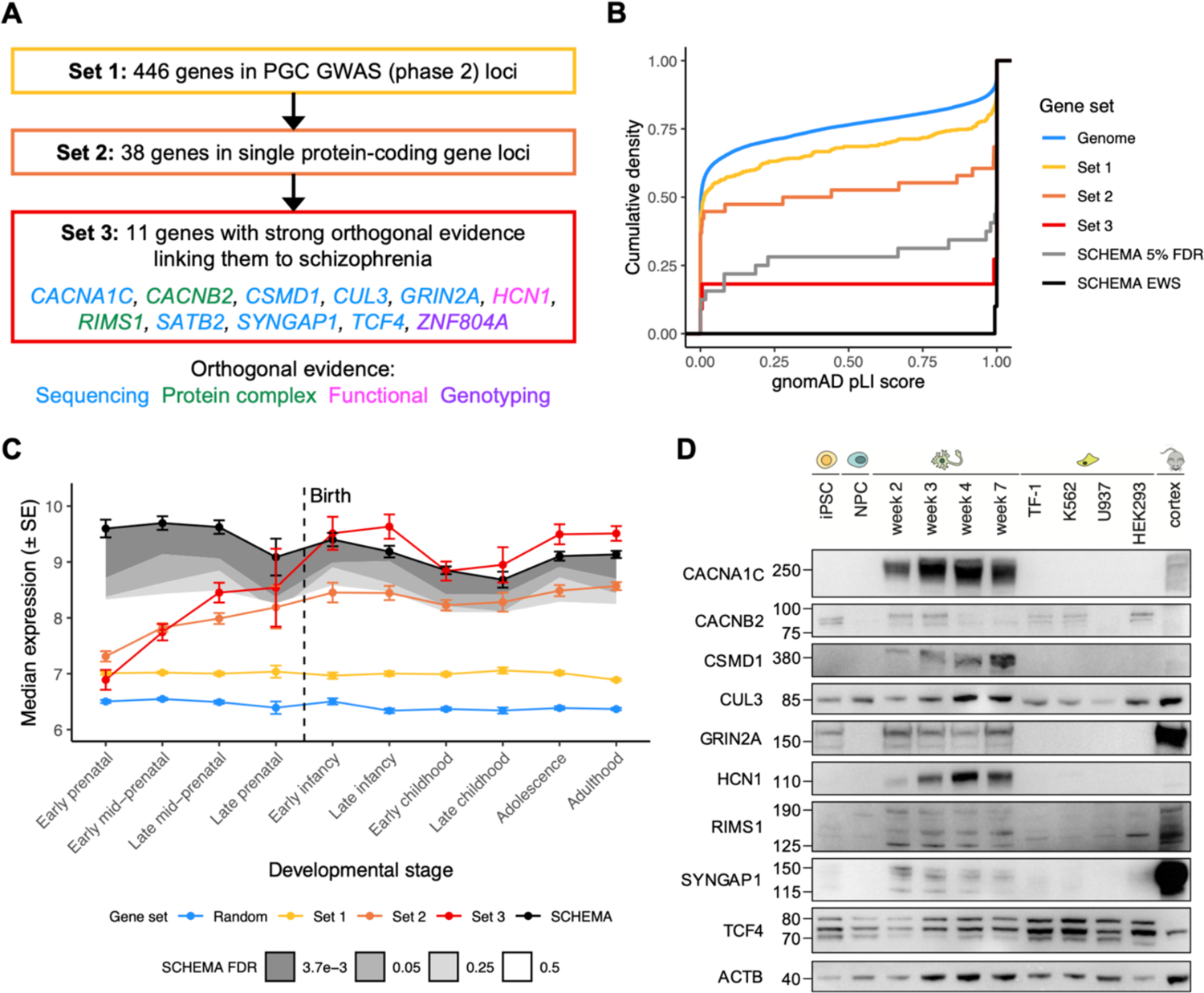
Selection of schizophrenia index genes and proteins for interactome experiments. **(A)** Three-step procedure to identify Sets 1-3 by refining schizophrenia GWAS [PGC phase 2] data, where Set 3 was defined as ‘index genes’ and used as the basis for downstream experiments. Set 3 genes are color-coded based on the type of orthogonal evidence supporting their involvement in neuropsychiatric or neurodevelopmental phenotypes. **(B)** Cumulative density of gnomAD pLI scores for different gene sets. ‘Genome’ indicates genes in the pLI dataset [excluding Sets 1-3]; ‘Sets 1-3’ indicate genes in Sets 1-3 with available pLI scores; ‘SCHEMA 5% FDR’ and ‘SCHEMA EWS’ indicate genes with FDR < 0.05 or 3.7e-3 [exome-wide significance] in the SCHEMA exome sequencing analysis, respectively. **(C)** Frontal cortex RNA expression of gene sets across ten developmental stages. Median expression and standard error [SE] of each gene set were derived from the BrainSpan exon microarray dataset. ‘Random’ indicates genes randomly sampled from the BrainSpan dataset; ‘Sets 1-3’ indicate genes in Sets 1-3 with available BrainSpan data; ‘SCHEMA’ indicates exome-wide significant genes from SCHEMA. Shaded regions indicate median expression of genes with FDR < 3.7e-3 [exome-wide significance], 0.05, 0.25, or 0.5 in SCHEMA with darker grey indicating greater significance. **(D)** Western blot analysis of index proteins in iPSCs, NPCs [at day 3 of differentiation], iNs [at weeks 2-7 of differentiation], three cancer cell lines [TF-1, K562, U937], HEK293 cells, and mouse cortex. SATB2 and ZNF804A are excluded from this panel due to lack of detectable expression in iNs.

Genes implicated in schizophrenia are under strong genetic selection and have elevated expression in the frontal cortex (*15*). Therefore, in order to assess the enrichment of schizophrenia risk genes in Sets 1-3, we compared their gnomAD (*16*) pLI scores (i.e., the probability of being loss-of-function intolerant, where genes under strong genetic selection have higher pLI scores) and BrainSpan (*17*) expression to schizophrenia risk genes from the recent Schizophrenia Exome Sequencing Meta-Analysis (SCHEMA) study (*15*). In terms of pLI scores, Sets 1-3 all have increasingly higher scores compared to other genes in the genome (one-tailed KS test *P* = 1.1e-3, 1.7e-3, and 1.3e-5 for Sets 1-3, respectively; **Fig. 1B** and **Data S2**). Set 3 scores are significantly higher than that of Set 2 (one-tailed KS test *P* = 5.8e-4), which in turn are higher than that of Set 1 (one-tailed KS test *P* = 0.019). Furthermore, Set 3 is under the same degree of genetic selection as the exome-wide significant (FDR < 3.7e-3) SCHEMA genes (two-tailed KS test *P* = 0.23). In terms of gene expression, the expression profile of Set 1 mirrors that of random genes during frontal cortical development up until adulthood; Set 2 has a postnatal expression profile that resembles SCHEMA genes with FDR of 0.25-0.5; and Set 3 has a postnatal expression profile that strongly mirrors the exome-wide significant SCHEMA genes (**Fig. 1C** and **Data S3**). While the lower prenatal expression of Set 3 compared to the SCHEMA genes may be reflecting different aspects of schizophrenia-related biology captured by common vs. rare variants, respectively (**Text S4**), the pLI scores and postnatal expression patterns generally support our three-step approach to gene selection based on refining GWAS data and indicate that Set 3 is enriched for bona fide schizophrenia risk genes. We proceeded to use Set 3 as the starting point of our experiments and refer to these 11 genes and their encoded proteins as ‘index genes’ and ‘index proteins’, respectively.

To study the expression patterns of the index proteins across different stages of iN maturation, we tested 58 commercially available antibodies and identified 31 with competency to detect the 11 index proteins (**Data S4**). We differentiated iPSCs to day three and weeks two, three, four, and seven, and confirmed protein expression of CACNA1C, CACNB2, CSMD1, CUL3, GRIN2A, HCN1, RIMS1, SYNGAP1, and TCF4 in neuron lysates by western blot (**Fig. 1D** and **Fig. S1**); SATB2 and ZNF804A lacked detectable neuronal expression or high-quality reagents and were excluded from further experiments. When index protein expression is compared between iNs, three cancer cell lines (i.e., TF-1, K562, and U937), HEK293 cells, and mouse cortex homogenate, CACNA1C, CSMD1, GRIN2A, HCN1, RIMS1, and SYNGAP1 display a neuron-specific expression profile in iNs compared to all other cell types.

Next, we tested 42 antibodies for their ability to immunoprecipitate (IP) the index proteins (**Data S4**) and were able to IP seven index proteins (CACNA1C, CACNB2, CUL3, HCN1, RIMS1, SYNGAP1, TCF4) in ∼seven billion iNs for subsequent mass spectrometry (MS) analyses. In total, we carried out 23 IP-MS experiments at five neuron differentiation time points (weeks one, two, three, four, and seven). We performed quality control (QC) and analyzed each experiment using Genoppi (*18*), calculating the log_2_ fold change (FC) and corresponding statistical significance for each protein identified in the index protein IPs compared to the controls, and then defining proteins with log_2_ FC > 0 and FDR ≤ 0.1 as the significant interactors of the index protein. We disregarded four IP experiments that did not meet our QC criteria (i.e., the log_2_ FC correlation between replicates is < 0.5 or the index protein is not positively enriched at FDR ≤ 0.1). The remaining 19 high-quality IPs of CACNA1C, CUL3, HCN1, RIMS1, SYNGAP1, and TCF4 have a median replicate log_2_ FC correlation of 0.87, with the six index proteins enriched at a median FDR of 8.2e-4 (**Fig. 2A,B, Fig. S2A,B, Data S5** and **S6**). In addition, we performed both experimental and computational analyses to confirm that, despite using an inclusive FC cutoff (i.e., log_2_ FC > 0) to define index protein interactors in the IPs, the interactors show no obvious quality differences across a range of different FCs (**Text S5**).

**Fig. 2.**
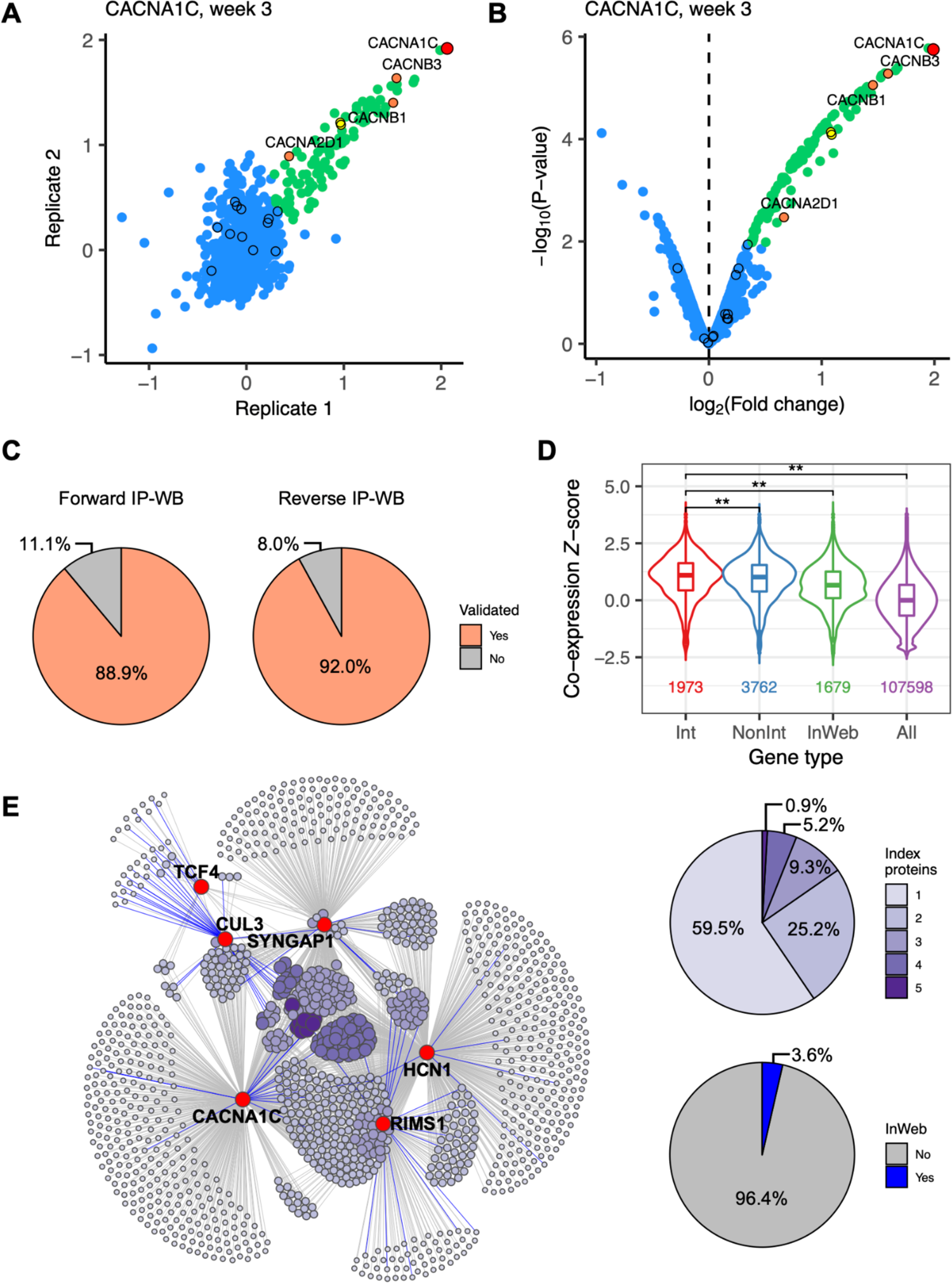
Cell-type-specific protein interactomes in cortical human neurons. **(A)** Scatter plot showing log_2_ FC correlation between replicate 1 [x-axis] and replicate 2 [y-axis] of an IP of CACNA1C at week 3 of neuron differentiation [Pearson’s *r* = 0.74]. **(B)** Volcano plot showing log_2_ FC [x-axis] and -log_10_ P-value [y-axis] of the CACNA1C IP from (A). For (A) and (B), the index protein [CACNA1C] is shown in red, significant interactors [log_2_ FC > 0 and FDR ≤ 0.1] in green, and non-interactors [i.e., other detected proteins] in blue. Known InWeb interactors are indicated by black border circles, with the subset that are significant in the IP highlighted in yellow [overlap *P* = 1.8e-2]. Calcium channel components [alpha, beta, and alpha2delta subunits] are in orange. **(C)** Validation rates of a subset of interactions tested in forward or reverse IPs followed by western blotting [IP-WB]. **(D)** Pairwise co-expression Z-scores between index genes and their interactors [Int], non-interactors [NonInt], known InWeb interactors [InWeb], and all protein-coding genes [All] derived from a spatial transcriptomic dataset in human dorsolateral prefrontal cortex. Double asterisks indicate significant difference in score distribution as calculated by a two-tailed Wilcoxon rank-sum test [*P* < 0.05/6, adjusting for 6 possible pairwise comparisons]. Number of gene pairs plotted for each gene type is indicated towards the bottom of the plot. **(E)** The combined interaction network of six index proteins resulting from 19 individual IPs. Index proteins and their interactors are indicated as red and purple nodes, respectively. Size and color of the interactor nodes scale with the number of index proteins linked to each interactor, with larger and darker nodes representing more recurrent interactors [distribution shown in upper right pie chart]. Edges represent protein interactions with colors indicating whether each interaction is known in InWeb [blue] or potentially novel [grey; distribution shown in lower right pie chart].

Importantly, our experimental design was purposely aimed at identifying large numbers of proteins by preserving non-stoichiometric interactions, rather than resolving direct interactions and core molecular complexes (**Text S6**). In previous work we have established that the validation rate of interactions identified using a similar IP-MS approach, cell model, and QC thresholds is 70-90% (*18*). In the current study, we were able to recapitulate these estimates using a two-pronged validation approach (**Fig. 2C**, **Fig. S3-S5**, and **Data S7**). First, we performed western blots of 45 interactors for the six index proteins upon repetition of the index protein IPs and validated 40 of the interactors (88.9% validation rate). In parallel, we also performed ‘reverse’ IPs using a panel of interactors as baits, and successfully detected the original index proteins in 23 out of 25 reverse IPs that showed bait enrichment (92.0% validation rate). Together these validation experiments indicate that the false positive rate in our data is in general agreement with the 10% FDR cutoff we applied to identify significant interactions.

Besides experimentally validating a subset of the index protein interactors, we also used published datasets to assess the biological validity of our IP-MS data. First, ten of the 19 IPs are further supported by the observation that they are enriched for known protein interactors derived from the InWeb_InBioMap (InWeb, hereafter) resource (*19*) (**Data S5**). As an example, in a CACNA1C IP performed in iNs at week 3 of differentiation, the significant interactors are enriched for known CACNA1C interactors in InWeb (*P* = 1.8e-2), including all known L-type calcium channel subunits: the extracellular CACNA2D1 and the intracellular CACNB1 and CACNB3 (*20*) (**Fig. 2A,B**). On the other hand, > 94% of the interactors are not found in InWeb nor in an IP of CACNA1C executed in mouse heart tissue with the same antibody (*21*). This example illustrates that our neuron-derived data capture both known and novel biology, which is expected given that existing protein-protein interaction datasets were mostly generated in non-neuronal context using different experimental methods (**Text S6**).

We also used brain co-expression data to systematically benchmark all index protein interactors in our data, reasoning that on average, transcripts of interacting proteins would be more likely to co-localize across tissues, cell types, and developmental time points (**Materials and Methods**). Indeed, we observed that the interactors usually have higher co-expression with the index proteins compared to the ‘non-interactors’ (i.e., non-significant proteins with log_2_ FC ≤ 0 or FDR > 0.1 in IP-MS), known interactors in InWeb, and all protein-coding genes in a spatial transcriptomic dataset derived from human dorsolateral prefrontal cortex (*22*) (**Fig. 2D**); similar trends were also observed in other expression datasets from human or mouse brains (*23, 24*) (**Fig. S2C**). This not only indicates that the interactions we identified in *in vitro* neurons represent biology found in complex brain tissues, but further suggests that they may be more enriched for gene relationships in the brain compared to proteins generally expressed in neurons (represented by the non-interactors) or interactions found in non-neuronal context (represented by the InWeb interactors). In summary, the experimental validation, InWeb overlap, and brain co-expression results all support the quality, reproducibility, and biological relevance of the interactome data we have generated for schizophrenia-related index proteins in human iNs.

When we compared multiple IPs of the same index protein across time points during neuronal maturation and between cell lines (for CACNA1C, HCN1, and SYNGAP1), we observed significant agreement in terms of the log_2_ FC correlation of all detected proteins (median correlation = 0.75; **Fig. S6A**,**C**,**E**). When we clustered the IPs based on the percentage of overlap between significant interactors, we found that IPs from the earlier vs. the later time points tend to fall into separate clusters, agreeing with known characteristics of maturing neurons generated using the NGN2 protocol (*14*) (**Fig. S6B**,**D**,**F** and **Data S8**). However, we also observed relatively high percentages of overlap that are statistically significant across all time points (median percentage = 72%). Overall, these results indicate that our IPs from different time points capture a large proportion of overlapping biology, which is not unexpected given that differentiating iPSCs start to express neuronal markers soon after forcing NGN2 overexpression. Based on these observations, we decided to explore the combined interaction network of each index protein across time points in downstream analyses.

We merged data from the 19 individual IPs to create nine additional consolidated datasets (**Data S9**). These datasets represent the combined interaction network of a single index protein across multiple experiments or time points (i.e., CACNA1C, HCN1, RIMS1 and SYNGAP1), the combined network of multiple index proteins at one time point (i.e., week 2, 3, 4, and 7), and the combined network of all six index proteins across all time points (i.e., ‘all combined’). The all combined network contains 1,238 interactors of the six index proteins and, similar to the CACNA1C IP highlighted above, > 96 % of the interactions in this network are potentially novel interactions not found in InWeb (**Fig. 2E** and **Data S5**). Genes encoding these interactors have relatively high expression in the frontal cortex throughout brain development similar to schizophrenia risk genes reaching 5% FDR in SCHEMA (**Fig. S7A**, **Data S3**, and **Text S4**). SynGO (*25*) gene set analysis also found the network to be enriched for genes involved in various biological processes in the synapse (**Fig. S7B** and **Data S10**). Overall, we successfully mapped the neuronal protein interactomes of six proteins that are transmembrane (CACNA1C, HCN1), cytosolic (CUL3, SYNGAP1), and involved in multiple neuronal signaling processes (RIMS1, SYNGAP1, TCF4). The resulting interaction networks are high-quality, include a high percentage of newly reported interactions, and span many areas of the cell biology of cortical excitatory neurons.

To test the networks for association to schizophrenia, we assessed the enrichment of common variant risk across the networks using PGC GWAS data containing schizophrenia cases and controls of European (EUR) or East Asian (EAS) ancestry (*3, 8*). For these analyses we created a more conservative version of the combined networks by excluding proteins that showed up as non-interactors in any of the source IPs that went into a corresponding network (see ‘stringent interactors’ defined in **Data S9**). In total, we performed genetic analyses for 11 interaction networks, including the nine combined networks and two individual IP networks for CUL3 and TCF4. Importantly, our analyses are conditional on the non-interactors detected in our IP experiments, meaning that we tested whether interactors in the different networks are enriched for genetic risk compared to other proteins expressed in iNs. If so, this would suggest that our networks are relevant to schizophrenia over and above the background proteome of the neuronal cell model.

Using MAGMA (*26*), we found that many of the interaction networks are indeed enriched for schizophrenia risk when conditioned on other proteins expressed in iNs. Notably, the enrichment is overall consistent across EUR and EAS ancestries (**Fig. 3A** and **Data S11**). We observed at least nominal significance (*P* < 0.05) in both ancestries for the all combined network, the week 4 network (includes IPs of CACNA1C, HCN1, RIMS1, and SYNGAP1), and the HCN1 network (includes IPs at weeks 2, 3, 4, and 7). Additionally, the CACNA1C network shows nominal significance in EUR and the same direction of effect in EAS; and vice versa for the week 2, week 7, and SYNGAP1 networks. At a Bonferroni-corrected P-value threshold (*P* < 0.05/22, adjusting for 11 networks and two ancestries), many of the networks remain significant, including the week 4 network in both EUR and EAS ancestries and the all combined, week 2, week 7, and HCN1 networks in EAS. We also performed a cross-ancestry meta-analysis, in which the all combined (nominal *P* = 1.0e-6, adj. *P =* 2.2e-5), week 2 (nominal *P* = 1.6e-3, adj. *P =* 0.035), week 4 (nominal *P* = 9.3e-6, adj. *P* = 2.0e-4), week 7 (nominal *P* = 1.3e-5, adj. *P* = 2.9e-4), and HCN1 (nominal *P* = 1.7e-5, adj. *P* = 3.7e-4) networks are all significant at the same Bonferroni-corrected threshold. In parallel, we further validated these findings using a genetic risk score enrichment analysis method that estimates the genetic risk on holdout samples not included in GWAS and therefore is less sensitive to outliers (**Materials and Methods**). Most of the findings were replicated, including the significant results for the all combined, week 4, and HCN1 networks in both ancestries and their meta-analysis (**Fig. S8B** and **Data S11**).

**Fig. 3.**
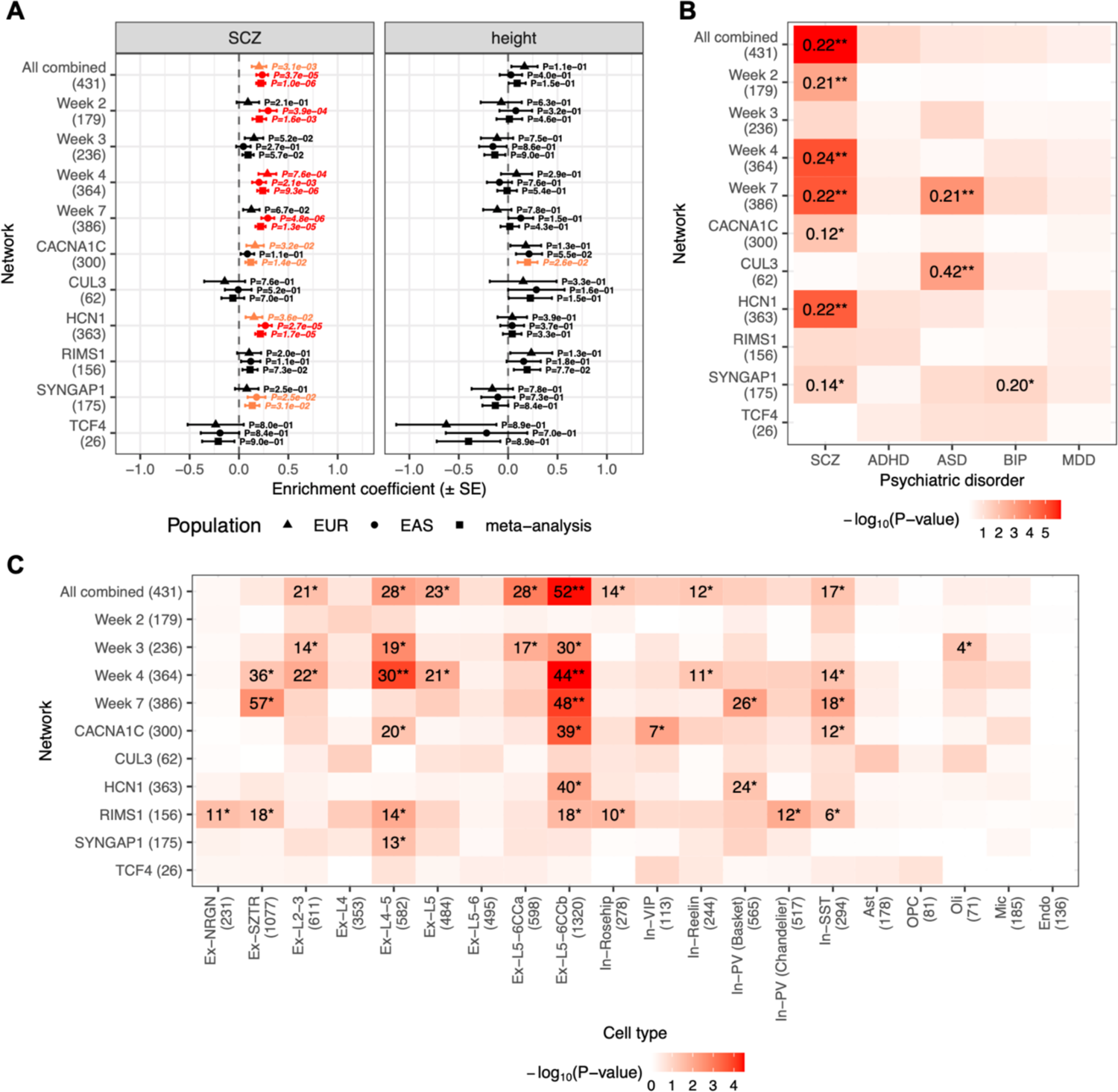
Enrichment of common variant risks and transcriptional perturbations in the index protein interactomes. Networks tested are the combined network of all IPs [All combined], the combined networks at each time point [Week 2 to Week 7], the combined networks for CACNA1C, HCN1, RIMS1, and SYNGAP1, and the individual IP networks for CUL3 and TCF4; the number of genes in each network is shown in parentheses on the y-axes. **(A)** Common variant enrichment of schizophrenia [SCZ] or height in Europeans [EUR], East Asians [EAS], or their meta-analysis. Enrichment coefficients, standard errors [SE], and P-values were calculated using MAGMA. Nominal [*P* < 0.05] or Bonferroni [*P* < 0.05/22, adjusting for 11 networks and two ancestries] significance is highlighted in orange or red, respectively. **(B)** Common variant enrichment of SCZ, attention deficit hyperactivity disorder [ADHD], autism spectrum disorders [ASD], bipolar disorder [BIP], or major depressive disorder [MDD] calculated using MAGMA. Cross-ancestry meta-analysis results are shown for SCZ; EUR ancestry results are shown for other disorders. Enrichment coefficients reaching nominal or Bonferroni [*P* < 0.05/22] significance are shown in the heat map followed by single or double asterisks, respectively. **(C)** Enrichment of cell-type-specific differentially expressed genes [DEGs] in the prefrontal cortex of schizophrenia patients compared to controls; the number of DEGs in each cell type is shown in parentheses on the x-axis. P-values were calculated using one-tailed hypergeometric tests. Gene counts in overlaps reaching nominal or Bonferroni [*P* < 0.05/220, adjusting for 11 networks and 20 cell types] significance are shown in the heat map followed by single or double asterisks, respectively.

To further explore whether the genetic risk enrichment we observed in the interaction networks is specific to schizophrenia, we repeated the same analyses using GWAS data of other psychiatric disorders, including attention deficit hyperactivity disorder (ADHD) (*27*), autism spectrum disorders (ASD) (*28*), bipolar disorder (BIP) (*29*), and major depressive disorder (MDD) (*30*), as well as height (*31, 32*) as a negative control (**Fig. 3A,B**, **Fig. S8A,B**, and **Data S11**). Across these phenotypes, we only observed robust enrichment for ASD in the CUL3 network and the week 7 network, which contains IP data of CUL3 and SYNGAP1, both of which have been previously linked to ASD (*33, 34*). Interestingly, despite the strong genetic correlation between schizophrenia and bipolar disorder (*29*), we only found nominally significant enrichment for BIP in the SYNGAP1 network. This lack of enrichment may be due to the fact that most of the index proteins are not BIP-specific risk proteins (i.e., only *CACNA1C* is in genome-wide significant loci of the BIP GWAS), or due to statistical power differences between the GWAS data for the two disorders. In any case, the networks that show the most robust enrichment for schizophrenia (i.e., the all combined, week 4, and HCN1 networks) do not show enrichment for the other phenotypes. This supports that most of the index protein interactors we identified in iNs are specifically concentrating common variant risk of schizophrenia.

Besides analyzing data from common variants, we also tested whether the interaction networks are enriched for rare variant risk of schizophrenia using gene-based association statistics from SCHEMA, and only observed nominal significance for the CUL3 network (**Fig. S8C** and **Data S11**). This is in contrast with the robust common variant enrichment results we observed for these networks, and repeating the analysis using larger sequencing datasets in the future will be needed to determine whether the lack of agreement is due to true biological differences between common and rare variants associated with schizophrenia or due to current power limitations in the genetic data. In parallel, we performed analogous analyses using association statistics from sequencing studies of developmental disorders (DD) (*35*) and ASD (*34*), as well as gnomAD pLI scores. At a Bonferroni-corrected P-value threshold (*P* < 0.05/11, adjusting for 11 networks), we found the CUL3 network to be enriched for DD genes, which is in agreement with previously implicated role of CUL3 in developmental delay (*33, 36*). In addition, the week 4 and RIMS1 networks are significantly enriched for genes with high pLI scores, indicating that some members of these networks are likely intolerant to loss-of-function mutations and may be involved in essential cell functions.

Next, we explored whether the interaction networks are enriched for brain-layer-specific transcriptional perturbations observed in patients with schizophrenia. We analyzed data from a recent single-cell RNA sequencing study (*12*), which identified differentially expressed genes (DEGs) in schizophrenia patients versus controls in 20 annotated cell types in the prefrontal cortex. Many of our networks show nominally significant overlaps with the cell-type-specific DEGs in neuronal cell types, which include both upper and deep layer excitatory neurons and inhibitory interneurons (**Fig. 3C** and **Data S12**). A population of layer 5/6 cortico-cortical projection neurons (‘Ex-L5-6CCb’) shows the most robust enrichment, reaching Bonferroni significance (*P* < 0.05/220, adjusting for 11 networks and 20 cell types) for the all combined (nominal *P* = 3.5e-5, adj. *P* = 7.7e-4), week 4 (nominal *P* = 3.2e-5, adj. *P* = 7.0e-4), and week 7 (nominal *P* = 9.5e-5, adj. *P* = 2.1e-3) networks. Subsequently, we separately analyzed the up- and down-regulated DEGs in this cell type, showing that the enrichment signals are strongly driven by the down-regulated DEGs (**Fig. S9** and **Data S12**). As DEGs in ‘Ex-L5-6CCb’ were also independently found to be enriched for genes implicated by schizophrenia GWAS (*12*), there is an intriguing convergence between our results and findings in patients that converge on deep layer cortical excitatory neurons as a relevant cell type for studying cellular networks involved in schizophrenia.

Functionally interpreting GWAS data to identify causal genes based on genome-wide significant SNPs is a major challenge in the field of genetics. Since our analyses indicate that the combined interaction network of all IPs is genetically and transcriptionally relevant in schizophrenia, we exploited this insight and used the network to prioritize genes in schizophrenia GWAS loci. We created a ‘social Manhattan plot’ by integrating our interaction data with the most recent PGC schizophrenia GWAS (phase 3) of 69K cases and 237K controls (*37*). We highlighted observed interactions between the index proteins and other proteins (locus proteins, hereafter) encoded by genes in the 270 reported genome-wide significant loci (**Fig. 4A** and **Data S13**). In total, we identified 114 locus proteins in 72 loci that are linked to ≥ 1 index protein in the social Manhattan plot. We further overlap these locus proteins with those prioritized by fine-mapping (FINEMAP) or eQTL co-localization (summary-based Mendelian randomization, SMR) analysis (*35*), pinpointing seven proteins that were also prioritized by FINEMAP (ACTR1B, EPN2, GABBR2, KIAA1549, MSI2, NEGR1, PDE4B) and three proteins that were also prioritized by SMR (SF3B1, PCDHA2, PCDHA8). In addition, our network was able to nominate candidate genes in 44 distinct loci that lack prioritization results from FINEMAP or SMR analysis.

**Fig. 4.**
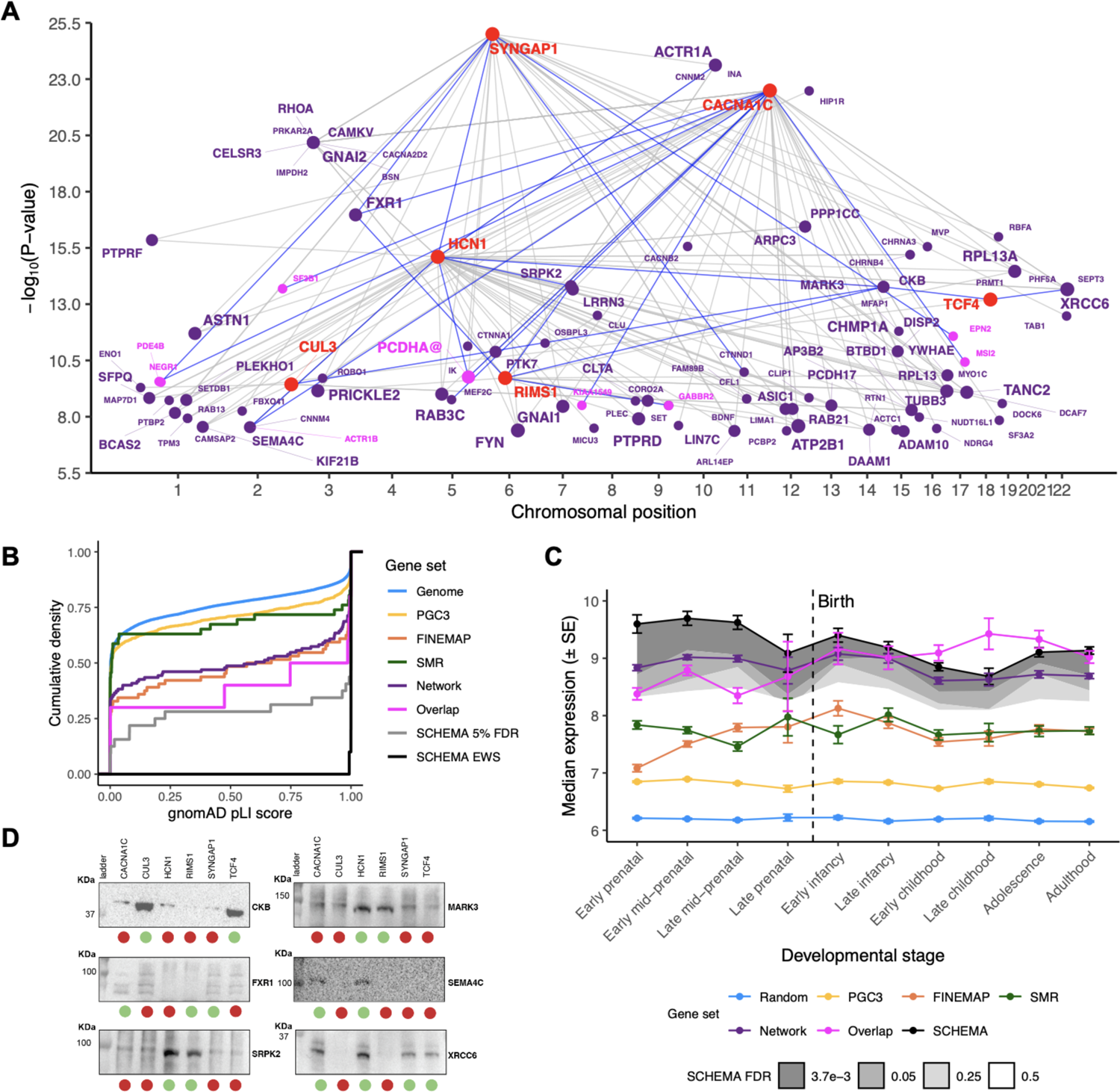
Prioritizing genes in schizophrenia GWAS loci using brain cell-type-specific interactome data. **(A)** Social Manhattan plot of genes encoding the index proteins [red] and their interactors [purple] in genome-wide significant loci in PGC schizophrenia GWAS [phase 3]. Size of the interactor nodes and their labels scale with the number of index genes linked to each interactor; those that were also prioritized by FINEMAP or SMR analysis are highlighted in magenta. Grey lines indicate observed protein-protein interactions in our data; interactions that have been validated by IP-WB are highlighted in blue. **(B)** Cumulative density of gnomAD pLI scores for different gene sets. ‘Genome’ indicates genes in the pLI dataset [excluding PGC3 genes]; ‘PGC3’ indicates genes in PGC GWAS [phase 3] loci; ‘FINEMAP’, ‘SMR’, and ‘Network’ indicate PGC3 genes prioritized by FINEMAP, SMR, or our interactome data, respectively; ‘Overlap’ indicates genes prioritized by all three approaches; ‘SCHEMA 5% FDR’ and ‘SCHEMA EWS’ indicate genes with FDR < 0.05 or 3.7e-3 [exome-wide significance] in SCHEMA, respectively. **(C)** Frontal cortex RNA expression of gene sets across ten developmental stages. Median expression and standard error [SE] of each gene set were derived from the BrainSpan exon microarray dataset. ‘Random’ indicates genes randomly sampled from the BrainSpan dataset; ‘PGC3’, ‘FINEMAP’, ‘SMR’, ‘Network’, and ‘Overlap’ indicate gene sets as described in (B); ‘SCHEMA’ indicates exome-wide significant genes from SCHEMA. Shaded regions indicate median expression of genes with FDR < 3.7e-3 [exome-wide significance], 0.05, 0.25, or 0.5 in SCHEMA with darker grey indicating greater significance. **(D)** Western blot analysis on independent IPs of index proteins [named on the top] to detect the presence of selected locus proteins [named on the side of each gel]. Green and red circles at the bottom represented whether the tested interaction was significant or non-significant by IP-MS, respectively. Each lane represents 10% of the IP material analyzed by IP-MS. L=ladder. Molecular weights are in KDa.

In order to compare our network prioritization approach to FINEMAP or SMR prioritization, we looked at the pLI score distributions and BrainSpan expression profiles of several gene sets, including all genes in the GWAS loci (PGC3), genes prioritized by our network (Network), FINEMAP, or SMR, and genes prioritized by all three approaches (Overlap). We observed that the Network and FINEMAP genes have comparable pLI scores (two-tailed KS test *P* = 0.92), and both sets have significantly higher scores compared to all PGC3 genes (one-tailed KS test *P* = 1.0e-6 and 3.6e-5, respectively), PGC3 genes in the same loci as the prioritized genes (one-tailed KS test *P* = 5.7e-7 and 3.9e-6, respectively), and the SMR genes (two-tailed KS test *P* = 3.8e-2 and 8.9e-3, respectively; **Fig. 4B** and **Data S2**). The Overlap genes have even higher pLI scores that are comparable to the SCHEMA genes with FDR < 0.05 (two-tailed KS test *P* = 0.27), although the differences between the Overlap genes and other genes in the Network, FINEMAP, or SMR supersets are not statistically significant (one-tailed KS test *P* = 0.10-0.60). When looking at gene expression throughout frontal cortical development, the Network, FINEMAP, SMR, and Overlap genes all have elevated expression compared to all PGC3 genes. In particular, the Network and Overlap genes have higher expression than the FINEMAP and SMR genes, and show postnatal expression profiles close to that of SCHEMA genes with FDR < 0.05 (**Fig. 4C** and **Data S3**).

Finally, we compared the Network genes to other PGC3 genes encoding proteins detected in our neuronal cell model during IP-MS (i.e., non-interactors in **Data S9**) and observed that these two gene sets have comparable pLI scores and BrainSpan expression (**Data S2** and **S3**). These results indicate that the proteome of the neuronal cell model may contribute to the pLI score and expression enrichment we observed for the Network genes. However, by first showing that our interaction network is significantly enriched for schizophrenia genetic risk and transcriptional perturbations over and above the non-interactors, and then showing that the network genes in GWAS loci have higher pLI scores and frontal cortical expression throughout development compared to other genes in the loci, we demonstrate how our interactome data can complement orthogonal methods such as statistical fine-mapping and eQTL co-localization analyses to nominate schizophrenia risk genes from GWAS data.

To demonstrate that many of the locus proteins we prioritized could be true interactors of the index proteins, we performed validation for selected interactors by repeating the index protein IPs followed by western blotting to detect the interactors, or vice versa, by executing reverse IPs for the interactors followed by western blotting to detect the index proteins. In total, we validated 24 interactions for 17 unique locus proteins using forward or reverse IPs, including eight proteins that were also prioritized by FINEMAP or SMR, as well as an interaction between two of the index proteins, HCN1 and SYNGAP1 (**Fig. 4A**, **Fig. S3-S5** and **Data S7**). In addition, we also performed western blots for several locus proteins on all six index protein IPs in parallel, showing that their detection patterns agree with their significant interactions with distinct index proteins in IP-MS (**Fig. 4D** and **Fig. S10**). These results illustrate the overall robustness and reproducibility of the interaction data we have generated and support the observed convergence between the index protein interactomes. Follow-up investigation on the locus proteins we have prioritized here can be informative for determining whether their corresponding genes are true schizophrenia risk genes responsible for the genetic signals observed in GWAS.

The last years have seen enormous progress in generating catalogs of genetic risk variants in schizophrenia, developing protocols to produce faithful stem cell models of human neurons relevant to disease, high-resolution proteomic approaches, and powerful computational frameworks to QC, validate, and integrate proteomic and genetic data. Here, we bring together these advances in a general framework to study cell-type-specific pathway relationships of schizophrenia risk genes in human cortical excitatory neurons. Our study showcases the vast, and nearly entirely untapped, potential for biological pathway discovery based on high-quality protein-protein interaction networks generated from human brain cell types. The robustness and biological validity of our interaction data are supported by the high (∼90%) validation rate in both forward and reverse IP experiments, and by the observation that interacting proteins are more likely to be co-expressed and co-localized in complex brain tissues. The interaction networks are further validated by the convergence of common risk variants from both European and East Asian patients, which implicates the networks in schizophrenia across populations and indicates that they will be a rich substrate for targeted follow-up mechanistic discoveries. This convergence is specific to schizophrenia, as the networks are generally not enriched for common variant risk of other psychiatric disorders. In addition, we integrated the networks with single-cell RNA sequencing data to pinpoint deep layer cortical excitatory neurons as a key cell type for schizophrenia, and with GWAS data to complement statistical fine-mapping and eQTL data for systematic interpretation of candidate genes in GWAS loci.

In our analyses, the HCN1 network emerges as a particularly promising lead for follow-up investigation. This network is enriched for schizophrenia common variant risk in both Europeans and East Asians, suggesting that perturbed signaling through the hyperpolarization-activated cyclic nucleotide-gated potassium channel, a heterotetrameric complex consisting of HCN1-4 (*38*), could play a role in schizophrenia across populations. Indeed, two HCN1 interactor genes we identified, *HCN4* and *AKAP11*, are also enriched for schizophrenia-associated protein-truncating variants (PTVs) in SCHEMA (FDR = 4.2e-3 and 1.3e-2, respectively) (*15*). In a recent meta-analysis of schizophrenia and bipolar disorder cases (*39*), *AKAP11* further emerged as an exome-wide significant (*P* = 2.8e-9) gene enriched for ultra-rare PTVs. PTVs are among the most interpretable genetic variants as their causal roles in disease are commonly linked to decreased gene function and expression. The observation that the HCN1 network is nominally enriched for down-regulated DEGs in layer 5/6 cortico-cortical projection neurons (‘Ex-L5-6CCb’) from schizophrenia patients (**Fig. S9**) also supports the hypothesis that members of the network may be involved in schizophrenia through loss-of-function or decreased expression.

Together, strong support from common variants and suggestive convergence of PTVs and transcriptional perturbations in patients with schizophrenia highlight a network involved in neuronal potassium signaling, which also contains numerous drug targets that can be explored in follow-up studies (**Fig. S11** and **Data S14**).

Another intriguing finding from our results is the recurrent interaction between CACNA1C and C4A (observed in four out of five CACNA1C IP-MS experiments; **Data S6**), suggesting that the L-type calcium channel may be a functional binding site of the complement cascade in synaptic pruning of the developing prefrontal cortex. This interaction is consistent with the emerging hypothesis that complement-mediated modulation of synapse stability or function contributes significantly to risk for schizophrenia (*40, 41*). In order to follow up on our findings, we performed substantial QC to identify suitable immunoreagents for IP of C4A (**Data S4**).

However, due to the complex post-translational modifications of C4A, we were unable to generate high-quality IP-MS data with the available antibodies in iNs. Interestingly, we also identified C3, another component of the complement cascade, as an interactor of CACNA1C in one of the CACNA1C IPs (**Data S6**). This interaction could be a perhaps better vantage point into the functional characterization of the interplay between synaptic biology and complement cascade, as it is more stable and therefore amenable to biochemical studies.

In the future, with larger genetic datasets to identify disease risk genes, wider availability of IP-competent immunoreagents, and the ability to create stem cell models of other brain cell types (e.g., inhibitory interneurons) at the scale required for systematic protein interactome experiments, we expect that the approach described here can be applied to uncover additional insights into the biology of schizophrenia and provide rich orthogonal information that is not captured by other approaches such as GWAS, exome sequencing, single-cell RNA sequencing, and whole-proteome analyses. More generally, our study establishes brain cell-type-specific protein interaction data as a key data type for studying psychiatric diseases, and provides an organizing framework to bring together genomic, transcriptomic, and proteomic data to model intracellular biochemical networks implicated in disease pathology. In fact, we have applied the same framework to study ASD risk genes derived from exome sequencing (co-submitted manuscript), showing that it strongly empowers interpretation of data generated from both common and rare variant genetics across two different groups of disorders. This framework can therefore contribute to laying the foundation for functional discovery and, eventually, rationally designed medicines in psychiatric diseases.

## Data Availability

Raw IP-MS data will be available through MassIVE upon publication of the manuscript.

## Acknowledgments

We thank Carole Manneville, Matthias Mueller, Katie Worringer, and Ajamete Kaykas at Novartis for sharing the hDFn cell line; Ellen Beauchamp, Andrew Guirguis, Rebecca Gorelov, and Zuzana Tothova for sharing cancer cell lines; Karl Clauser for managing and uploading the proteomics data; and Steve Hyman, Mark Daly, and Ben Neale for insightful scientific discussions.

## Author contributions

EN, NP, WC, JMS, GP, JR, JMM and JCB carried out tissue culture, tested antibodies for WB/IP experiments and ran WBs. EN, NP, WC, JMS and BT executed IP experiments. SBE, BT, CRS, AMA, MS and JJ ran MS experiments and analysis. SAC supervised the MS experiments. AK and EM analyzed and consolidated IP-MS data. YHH, KT and TL consolidated IP-MS data and ran BrainSpan analysis. RL performed common variant enrichment analyses. JKTC performed comparisons of IP-MS data. YHH performed the rest of the analyses. TS, TG and HH designed genetic risk enrichment analyses. NF supervised and managed the study. KCE and KL designed and supervised the study. KL initiated and led the study. YHH, EN, RL, KT, NF, HH, KCE and KL wrote the manuscript with input from co-authors.

## Data availability

Raw IP-MS data will be available through MassIVE upon publication of the manuscript.

## Supplementary Materials

Materials and Methods Text S1 to S6

Figs. S1 to S11

Data S1 to S14 References (42–71)

