## Supplementary Materials for "Using brain cell-type-specific protein interactomes to interpret genetic data in schizophrenia"

**This PDF file includes:**

Materials and Methods

Text S1 to S6

Figs. S1 to S11

Captions for Data S1 to S14

**Other Supplementary Materials for this manuscript include the following:**

Data S1 to S14 (Excel format)

### Materials and Methods

Refining genetic data to identify index genes

*Three-step procedure for selecting index genes:* Starting with 125 independent autosomal SNPs that reached genome-wide significance in the combined discovery-replication meta-analysis of the PGC schizophrenia GWAS (phase 2) (*3*), Ricopili (*42*) (https://data.broadinstitute.org/mpg/ricopili/) was used to map the 124 non-MHC (major histocompatibility complex) region SNPs to 445 genes (Set 1; **Data S1**) in linkage disequilibrium (LD) loci, whose boundaries were defined by SNPs in LD ($r^2 > 0.6$) with the index SNPs, ± 50kb on either end. Next, we excluded SNPs in intergenic regions or in loci with multiple protein-coding genes, resulting in a list of 40 SNPs pointing to 37 unique protein-coding genes (Set 2) in single-gene loci. We further refined the 37 genes into a set of 10 genes (Set 3) based on strong orthogonal evidence supporting their involvement in psychiatric diseases (see below). We also included *SYNGAP1* in the MHC region (which was excluded from the SNP-to-gene mapping process due to its LD complexity) in all 3 sets based on strong orthogonal evidence. In total, we identified 11 high-confidence schizophrenia risk genes and considered their encoded proteins as index proteins in the proteomic experiments.

*Orthogonal evidence for the index genes:* The index genes were selected based on several types of orthogonal genetic or functional evidence, including: i] high-density genotyping experiments of individual genes (*ZNF804A*) (*43*), ii] sequencing or copy number variant studies linking genes to autism spectrum disorders or neurodevelopmental delay (*CACNA1C*, *CUL3*, *CSMD1, GRIN2A*, *SATB2 SYNGAP1*, *TCF4*) (*33*, *44–51*), iii] strong functional evidence supporting that the gene in question is causal (*HCN1*) (*52*), and iv] multiple subunits from the same protein complex are encoded by genes in different single-gene loci linked to psychiatric diseases (*CACNA1C*, *CACNB2*, *RIMS1*) (*3*, *53*, *54*). In addition, we annotated the 10 genes prioritized from single-gene GWAS loci using Hi-C chromatin interaction data from the developing human brain (*55*), which may point to long-range regulation of genes outside of the loci in the context of schizophrenia; only three of these genes (*CUL3, RIMS1, SATB2*) lie in loci that exclusively interacted with long-range genes outside of the loci (**Data S1**).

pLI score enrichment analysis

We extracted pLI scores from the gnomAD (*16*) (v2.1.1) 'pLoF Metrics by Gene TSV' dataset. We performed one-tailed Kolmogorov-Smirnov (KS) tests to assess whether various gene sets are enriched for high pLI scores compared to other genes in the genome. The tested gene sets include: i] SCHEMA (*15*) genes with FDR < 3.7e-3 (exome-wide significance) and 0.5, ii] Sets 1-3 defined by our index gene selection steps, and iii] genes in PGC schizophrenia GWAS (phase 3) loci (*37*) and subsets of these genes prioritized by FINEMAP/SMR analysis and/or our interaction data (see **Social Manhattan plot**). In addition, we performed one-tailed KS tests to compare the pLI scores of several gene sets that are subsets and supersets of each other, and two-tailed KS tests to compare a few disjoint sets. **Data S2** summarizes all the comparisons performed. Finally, we also performed one-tailed KS tests to assess whether the stringent interactors in our interaction networks (**Data S9**) have enriched pLI scores compared to the non-interactors linked to each network.

BrainSpan expression profiles

We obtained gene expression data in four distinct parts of the frontal cortex (dorsolateral prefrontal cortex [DFC], medial prefrontal cortex [MFC], ventrolateral prefrontal cortex [VFC], orbital frontal cortex [OFC]) across 10 developmental stages from the BrainSpan (https://www.brainspan.org) exon microarray dataset (*17*). At each developmental stage, we calculated the median and standard error of the expression values for various genes or gene sets including: i] SCHEMA genes with FDR < 3.7e-3, 0.05, 0.25, and 0.5, ii] Sets 1-3 defined by our index gene selection steps, iii] index and interactor genes in our combined network of all IPs (**Data S9**), iv] genes in PGC schizophrenia GWAS (phase 3) loci and subsets of these genes prioritized by FINEMAP/SMR analysis and/or our interaction data (see **Social Manhattan plot**), and v] random genes sampled from the BrainSpan dataset for comparison against the other gene sets. **Data S3** provides more detailed summary statistics for all gene sets.

iN differentiation

Glutamatergic patterned induced neurons (iNs) were differentiated from male stem cells by conditional expression of the neuralizing transcription factor NGN2 as previously described (*14*), with the exception that N2 media was used instead of KSR media during days 0 to 3. iNs were re-passaged at day 3 of differentiation (i.e., 40,000 cells/cm2) on Geltrex (Thermo Scientific #A1413202) coated plates. In order to remove remaining proliferating cells, fluoro-deoxyuridine (Bioworld 40690016-2) was added to cell cultures at 10 µM on day 6 of differentiation. iNs used in a subset of the CACNA1C, HCN1, and TCF4 IP-MS experiments (**Data S5**) were differentiated from human embryonic stem cells WA01 (H1, NIH registration no. 0043) transduced with lentiviruses carrying TetO::Ngn2-Puro and reverse tetracycline-controlled transactivator (rtTA). All other IP-MS experiments used iNs generated from a clonally selected induced PSC line (iPS hDFn 83/22 iNgn2#9 [iPS3]) with TetO::Ngn2 and rtTA delivered via PiggyBAC (*14*).

Cell lines and tissue culture

*iNs:* iNs collected at time points under one week of differentiation were dissociated from plates with Accutase (Innovative Cell Technologies AT104-500) at 37°C for 5-10 min. iNs collected at time points beyond two weeks of differentiation were collected by removing media, adding 1x PBS, and swirling the plates until the neurons detached. Dissociated or detached cells were collected in 15 mL or 50 mL Falcon tubes depending on the number of cells and washed 3 times. Each wash consisted of centrifugation at 300 RCF for 3-5 min, discarding of the supernatant, and resuspension of the cell pellet in 1x PBS. After the final wash, the sample was centrifuged at 300 RCF for 3-5 min, supernatant was discarded, and cells were either lysed or flash frozen in liquid nitrogen and stored at -80°C. Lysis was performed according to manufacturer's protocol using Pierce IP lysis buffer (Thermo Scientific #87788) with 1x Halt Protease and Phosphatase cocktail inhibitor (Thermo Scientific #7844). Protein concentration of the lysate was quantified using BCA Protein assay (Thermo Scientific #23227). When not used for IP-MS experiments, the lysate was aliquoted in low-bind microfuge tubes (Axygen Scientific #MCT-175-L-C), flash frozen and stored at -80°C.

*Cancer and HEK cells:* We used the following cell lines as controls in western blots: HEK-293 (ATCC CRL-1573), a human embryonic kidney cell line; TF-1 (ATCC CRL-2003), a human erythroleukemia cell line; K-562 (ATCC CCL-243), a human myelogenous leukemia cell line; and U-937 (ATCC CRL-1593.2), a histiocytic lymphoma cell line. All cell lines were grown on uncoated plates (Corning) according to vendor recommendations. Media was changed every 3 days and cells were passaged when reaching 70% confluency. All cell lines were incubated at 37 °C, 5% $CO_2$. TrypLE (Thermo Scientific) was used to detach cells from plates.

Mouse cortex sample preparation

Mouse cortices were isolated from p0 pups of C57BL/6 background, cut into small pieces, and flash frozen. Lysis was performed by adding Pierce IP lysis buffer to frozen pieces and immediately applying a handheld homogenizer (VWR pellet mixer #47747-370). All subsequent steps were the same as those for iNs.

Western blotting

For western blots, frozen lysates were thawed on ice, diluted to desired concentration in 1x PBS, and brought to 1x LDS using 4x LDS stock. Samples were denatured at various conditions based on what we identified to work best for each protein of interest. Prepared samples were run on NuPAGE 1.5mm 3-7% Tris-Acetate or 1mm 4-12% Bis-Tris gels (Thermo Scientific). Transfer was onto a nitrocellulose membrane using XCell wet transfer (Invitrogen), iBlot2 (Thermo Scientific), or Trans-Blot Turbo (BioRad). Membranes were blocked with 5% BSA or 5% milk diluted in TBST, incubated with primary antibody diluted in matching blocking buffer for at least 12h at 4°C, and washed 3 x 10 min in TBST. Membranes were incubated with HRP-conjugated secondary antibody diluted in 5% milk for 1h at RT, washed 3 x 10 min in TBST, developed with enhanced chemiluminescent substrate (Thermo Scientific #34095), and imaged on ChemiDoc MP (BioRad). In 'forward' IP western validation, the immunoprecipitate of the index protein was loaded into the gel and incubated with the primary antibody of an interactor protein to determine if the interactor could be found in the immunoprecipitate; in 'reverse' IP western validation, the immunoprecipitate of an interactor was loaded into the gel and incubated with the primary antibody of the index protein to determine if the index protein could be found in the immunoprecipitate. Western blot antibodies used for each protein of interest are listed in **Data S4**.

Immunoprecipitations

For immunoprecipitations (IPs) followed by western blots or Coomassie stain analysis, either fresh or previously frozen lysates were used. For IPs followed by mass spectrometry (MS) only fresh lysates were used. On day 1, the needed amounts of lysate and IP antibody were added to a 1.7 mL Axygen MaxyClear tube (MCT-175-L-C), then brought to a final volume of 1.3 mL with Pierce IP lysis buffer. Tubes were then rotated at 4°C overnight for 14-18 h. On day 2, the needed amount of beads per IP were apportioned to separate tubes, washed twice in cold lysis buffer, resuspended in 200 µL lysis buffer per tube, added to corresponding tubes, and rotated at 4°C for 2-4 h. After incubation, tubes were placed on ice, and beads were first washed once in 1mL cold lysis buffer and then twice in 1x cold PBS. To remove supernatants in between the steps, magnetic beads were placed on a magnetic rack on ice and agarose beads were spun at 2,000 x g for 3 min. Supernatants after overnight rotation and each wash step were collected for western blot analysis of IP quality. After the third wash, supernatant was removed, and beads were resuspended in 50 µL of PBS if the IP was to be processed by MS and 40 µL of PBS if the IP was to be processed by western blot or Coomassie stain. Samples were flash frozen in liquid nitrogen and stored at -80°C until further use. All IP experiments sent for MS consisted of 4 IP samples performed on cells from the same differentiation batch: 2 replicate experimental IPs using an antibody against the protein of interest (i.e., an index protein) and 2 replicate control IPs using a control IgG antibody. IP antibodies used for each protein of interest are listed in **Data S4**.

Immunoprecipitations using V5 epitope tagged TCF4

For immunoprecipitation of TCF4, an ORF of the TCF4 isoform A with a c-terminal V5-tag was acquired from the Broad Institute Genetic Perturbation Platform, and lentiviruses were generated from the construct by ALSTEM Inc. Lentiviruses were delivered to iN cultures at a multiplicity of infection (MOI) of 4 at day 3 of differentiation during re-passaging, and mock-transduced cells were used as controls. Cells were collected at day 6 of differentiation by washing 3 times with PBS, followed by lysis and scraping on the plate. Lysates were then processed and quantified as usual. For IP, the needed amount of lysate was added to a microfuge tube and brought to a final volume of 1.3 mL with Pierce IP lysis buffer, then incubated overnight with anti-v5 antibody directly coupled to magnetic beads (MBL International, M167-11). The remaining protocol is the same as used for the other IPs.

Mass spectrometry

*Sample preparation:* Proteins were digested on beads using 90 µl of digestion buffer (2 M urea / 50 mM Tris buffer with 1 mM DTT and 5 µg/mL Trypsin) for 1 hr, shaking at 1000 rpm. The suspension was then transferred to a new tube, and the beads were washed twice with 60 µL of wash buffer (2 M urea / 50 mM Tris buffer). The wash buffer was added to the suspension with digestion. The digestion and wash process was repeated a second time pooling the suspensions with the suspensions from the first round. The pooled solution was reduced using 4 mM DTT for 30 min at 25°C shaking at 1000 rpm. The proteins were then alkylated using 10 mM iodoacetamide and incubating for 45 min at 25°C shaking at 1000 rpm and protected from light. Proteins were then digested with 0.5 µg of trypsin overnight at 25°C shaking at 700 rpm. The next day proteins were quenched using 40 µL of 10% formic acid and desalted using an Oasis Cartridge. Samples were vacuum dried and labeled with iTRAQ4 (Sciex Inc.) or TMT10 (Thermo Scientific) kits. Each iTRAQ 4-plex consisted of 2 replicate experimental IPs using an antibody against the protein of interest (i.e., an index protein), and 2 replicate control IPs using a control IgG antibody. The specific iTRAQ labels for each replicate pair are indicated in **Data S5**. For 'RIMS1_wk4_2', a TMT 10-plex experiment was conducted with antibodies from Santa Cruz Biotechnology (SC; 128C, 129N), Proteintech Group (PT; 126, 127N), and Synaptic Systems (SYSY; 129C, 130N). Two sets of replicate control IPs were included for PT (127C, 128N) and SC/SYSY (130C, 131). Only the results from the SC antibody met our quality control metrics and were reported.

*Liquid chromatography-tandem mass spectrometry (LC-MS/MS):* Reconstituted peptides were separated on an online nanoflow EASY-nLC 1000 UHPLC system (Thermo Scientific) and analyzed on a benchtop Orbitrap Q Exactive Plus mass spectrometer (Thermo Scientific). The peptide samples were injected onto a capillary column (Picofrit with 10 µm tip opening / 75 µm diameter, New Objective, PF360-75-10-N-5) packed in-house with 20 cm C18 silica material (1.9 µm ReproSil-Pur C18-AQ medium, Dr. Maisch GmbH, r119.aq). The UHPLC setup was connected with a custom-fit microadapting tee (360 µm, IDEX Health & Science, UH-753), and capillary columns were heated to 50 °C in column heater sleeves (Phoenix-ST) to reduce backpressure during UHPLC separation. Injected peptides were separated at a flow rate of 200 nL/min with a linear 150 min gradient from 94% solvent A (3% acetonitrile, 0.1% formic acid) to 35% solvent B (90% acetonitrile, 0.1% formic acid), followed by a linear 8 min gradient from 35% solvent B to 60% solvent B and a 3 min ramp to 90% B. The Q Exactive instrument was operated in the data-dependent mode acquiring HCD MS/MS scans (R=17,500 for iTRAQ4, or R=35,000 for TMT10) after each MS1 scan (R=70,000) on the 12 most abundant ions using an

MS1 ion target of 3x106 ions and an MS2 target of 5x104 ions. The maximum ion time utilized for the MS/MS scans was 120 ms; the HCD-normalized collision energy was set to 28 for iTRAQ4 or 31 for TMT10; the dynamic exclusion time was set to 20s, and the peptide match and isotope exclusion functions were enabled.

IP-MS data analysis

*Spectrum Mill analysis:* All mass spectra were processed using the Spectrum Mill software package (v7.0; https://proteomics.broadinstitute.org). For peptide identification, MS/MS spectra were searched against a sequence database for the Uniprot human reference proteome, including isoforms, with a set of common laboratory contaminant proteins appended (2017: 65,068 entries, or 2014: 59,079 entries) as indicated in **Data S5**. Search parameters included: ESI-QEXACTIVE-HCD scoring parameters, trypsin enzyme specificity with a maximum of two missed cleavages, 40% minimum matched peak intensity, ± 20 ppm precursor mass tolerance, ± 20 ppm product mass tolerance. Carbamidomethylation of cysteines and iTRAQ4 or TMT10 full labeling of lysines and peptide n-termini were set as fixed modifications. Allowed variable modifications were oxidation of methionine (M), acetyl (ProtN-term), and deamidated (N), with a precursor MH+ shift range of -18 to 64 Da. Identities interpreted for individual spectra were automatically designated as valid by optimizing score and delta rank1-rank2 score thresholds separately for each precursor charge state in each LC-MS/MS while allowing a maximum target-decoy-based false discovery rate (FDR) of 1.0% at the spectrum level. Identified peptides were organized into protein groups and subgroups (isoforms and family members) with Spectrum Mill's subgroup specific option enabled, so that peptides shared between subgroups are ignored for quantitation. TMT10 reporter ion intensities were corrected for isotopic impurities using Spectrum Mill's afRICA correction method and correction factors obtained from the reagent manufacturer's certificate of analysis. For quantitation at the peptide spectrum match level, $log_2$ fold change (FC) reporter ion intensity ratios were calculated for each IP replicate. To obtain protein-level $log_2$ FC values between index protein vs. control for each IP replicate, the median ratio was calculated from all peptide spectrum matches of subgroup specific peptides assigned to each protein subgroup.

*Genoppi analysis:* For each IP-MS experiment, starting with the protein-level quantification report generated by Spectrum Mill, we applied median normalization to protein $log_2$ FC values between index protein vs. control IPs for each replicate, and then performed downstream analyses using Genoppi (*18*) (v1.0). Additional data processing was performed for two experiments pre-Genoppi: for 'CACNA1C_wk7' the $log_2$ FC values for one replicate was right-shifted by a small value (0.005) to facilitate direct comparisons with all other volcano plots without changing the results; for 'TCF4_wk1' additional contaminant proteins found in the CRAPome (*56*) database were removed. The Genoppi analyses include: i] calculating Pearson's correlation of $log_2$ FC values between IP replicates, ii] calculating average $log_2$ FC, and corresponding P-value and FDR, for each protein across IP replicates using a two-tailed one-sample moderated t-test from the limma (*57*) R package, iii] identifying statistically significant ($log_2$ FC $> 0$ and FDR $\leq 0.1$) index protein interactors (i.e., proteins with significantly higher abundance in the index protein IPs compared to the controls), iv] defining other non-significant ($log_2$ FC $\leq 0$ or FDR $> 0.1$) proteins to be the 'non-interactors' (to serve as proxy for background proteome in enrichment analyses), and v] assessing overlap enrichment between the identified interactors and known interactors from InWeb_InBioMap (*19*) (compared to the non-interactors) using a one-tailed hypergeometric test. We performed quality control of each IP-MS experiment using two criteria: i] the $\log_2$ FC correlation between replicates must be > 0.5, and ii] the index protein itself must be significant ($\log_2$ FC > 0 and FDR ≤ 0.1). Experiments that failed to meet these criteria were excluded from further analysis; those that passed QC are summarized in **Data S5**, with their analysis results provided in **Data S6**.

Co-expression analysis

We assessed pairwise co-expression between each index gene and all other protein-coding genes using data from four independent studies, including: i] Stickels *et al.* (*23*): spatial transcriptomics in mouse neocortex (Puck_190921_19.digital_expression.txt.gz retrieved from: https://singlecell.broadinstitute.org/single_cell/study/SCP815/highly-sensitive-spatial-transcriptomics-at-near-cellular-resolution-with-slide-seqv2#study-download), ii] Maynard *et al.* (*22*): spatial transcriptomics in human dorsolateral prefrontal cortex (count matrix retrieved from spatialLIBD R package: https://github.com/LieberInstitute/HumanPilot), iii] Velmeshev *et al.* (*24*): single-cell RNA-seq in human cortex (rawMatrix.zip retrieved from: https://cells.ucsc.edu/?ds=autism), and iv] BrainSpan: bulk RNA-seq across human brain regions and developmental stages (“RNA-Seq Gencode v10 summarized to genes” dataset retrieved from: https://www.brainspan.org/static/download.html). We used two different methods for estimating co-expression to account for different statistical properties in these datasets. For the spatial transcriptomic datasets with very sparse gene expression matrices, we reasoned that the binary presence/absence of genes across physical locations would be the most informative, and therefore performed one-tailed Fisher’s exact tests to calculate the significance of co-occurrence for each gene pair across locations. For the single-cell RNA-seq and BrainSpan datasets, we calculated a proportionality metric, ρ, for each gene pair using the propr R package (*58*) (v4.2.6);

this metric is analogous to conventional correlation measures but has been shown to be better at capturing functional associations between genes in RNA-seq data (*59*). After calculating either the Fisher's exact P-values or the proportionality ρ values for all gene pairs involving each index gene, we then performed rank-based inverse normal transformation to convert the values into co-expression Z-scores (where a positive score indicates that a gene has higher than average co-expression with the index gene compared to the rest of the genome).

We performed two-tailed Wilcoxon rank-sum tests to assess if the co-expression Z-scores between index genes and their interactors are significantly different from the scores between the index genes and other gene groups including: i] non-interactors detected in IP-MS, ii] known interactors from InWeb_InBioMap (*19*), and iii] all protein-coding genes.

<u>Consolidating IP-MS datasets into interaction networks</u>

We consolidated IP-MS datasets for the same index proteins into index-protein-specific networks; datasets derived from the same time points into time point-specific networks; and all datasets into an all combined network. For each network, we defined 'interactors' as proteins that show up as significant interactors in ≥ 1 source IPs contributing to the network, and the matching 'non-interactors' as proteins that show up as non-interactors in ≥ 1 source IPs but never as interactors in these IPs. In addition, we defined 'stringent interactors' as interactors that never show up as non-interactors in the source IPs. Finally, we removed proteins whose HGNC gene symbols could not be mapped to Ensembl (*60*) (GRCh37.p13) genomic positions and the index proteins for the source IPs from all 3 lists to facilitate downstream enrichment analyses. **Data S9** lists the interactors, non-interactors, and stringent interactors associated with each network.

SynGO gene set analysis

SynGO analysis of interactor genes in the all combined network was performed using the SynGO web browser (*25*) (dataset version: 20210225; https://syngoportal.org). The SynGO 'Biological Processes' annotations and the 'brain expressed' background set were used in the analysis.

Common variant enrichment analysis using MAGMA

To perform common variant risk enrichment analysis for each interaction network using MAGMA (*26*) (v1.09b), PGC GWAS summary statistics (https://www.med.unc.edu/pgc/download-results/) were obtained for schizophrenia (33,640 cases and 43,456 controls of EUR ancestry (*3*); 22,778 cases and 35,362 controls of EAS ancestry (*8*) ), ADHD (19,099 cases and 34,194 controls of EUR ancestry (*27*)), ASD (18,382 cases and 27,969 controls of EUR ancestry (*28*)), BIP (41,917 cases and 371,549 controls of EUR ancestry (*29*)), and MDD (170,756 cases and 329,443 controls of EUR ancestry (*30*)). GWAS summary statistics for height were obtained from the Neale Lab UK Biobank GWAS (round 2; 361,194 individuals of EUR ancestry (*31*); https://www.nealelab.is/uk-biobank/) and Biobank Japan (159,095 individuals of EAS ancestry (*32*); http://jenger.riken.jp/en/result). For each GWAS dataset, we first annotated variants to genes using the Human reference genome (GRCh37/hg19) with a flanking gene region of ± 250kb. Variants on sex chromosomes, with minor allele frequency (MAF) ≤ 0.05 in the study, or within the MHC region (chr6:22.5M-33.5M) were excluded from analysis. Next, gene-based P-values were computed using the SNP-wise Mean model in MAGMA. LD was estimated from the 1000 Genomes Project (*61*) phase 3 EUR or EAS panel to match the ancestry of each study. Competitive tests were used for gene-set comparison analysis between stringent interactors and non-interactors in each interaction network. Specifically, a linear regression model was built by MAGMA to test if genes within the interactor gene-set are more strongly associated with the phenotype of interest compared to the non-interactor gene-set. Inverse-variance weighted fixed-effect meta-analysis (*62*) was used to combine results across EUR and EAS ancestries for schizophrenia and height, and a one-tailed P-value was calculated from the meta-analyzed Z-score.

Common variant enrichment analysis using the GRS method

To complement the MAGMA analysis, we used an alternative genetic risk score (GRS) method and individual-level genotypes from the schizophrenia GWAS study cohorts (24,764 cases and 30,655 controls of EUR ancestry; 8,960 cases and 8,284 controls of EAS ancestry) to assess genetic risk enrichment in the interaction networks. IRB approvals for accessing the individual-level data were obtained from the PGC and Stanley Global Asia Initiatives (**Text S3**). For each study cohort, we removed variants in the MHC region and variants with MAF $\leq 0.05$ or low imputation quality (INFO $\leq 0.8$), and then mapped the remaining variants to the stringent interactors or non-interactors associated with each network with a flanking gene region of ± 250kb. Next, we used PLINK (*63*) (v1.9) and the in-sample LD to clump variants within the interactor or non-interactor gene-sets into independent association signals. The clumping was performed with a window of 250kb, $R^2 > 0.2$, and a P-value threshold of 1. The out-of-sample GRS for each individual and each gene-set was then calculated as $\sum \log(OR) * G$, in which $OR$ is the estimated odds ratio from the leave-one-out GWAS meta-analysis for the clumped index variants within the gene-set and $G$ is the genotype dosage. To test if the stringent interactors in a network contribute more genetic risk compared to the non-interactors, we fit a linear regression model: $GRS_{ij} = P_i + I_j + P_i * I_j + Cov_i$, where $i$ and $j$ denote individual index and gene index, respectively, $P$ is the case/control status, $I$ is interactor/non-interactor status, $P * I$ is an interaction term, and $Cov$ includes 10 genetic principal components. We tested whether the $P * I$ interaction term is significantly larger than zero, which captures whether the difference of GRS between cases and controls calculated in the interactor gene-set is significantly larger than the non-interactor gene-set. Inverse-variance weighted fixed-effect meta-analysis was used to combine the test statistics for the interaction term across study cohorts and ancestries, and a one-tailed P-value was calculated from the meta-analyzed Z-score. Finally, as a negative control, we performed an analogous GRS analysis for height; in this case, the GRS was calculated as $\sum \log(beta) * G$, where *beta* is the effect size estimate from the EUR or EAS height GWAS for clumped index variants.

Rare variant enrichment analysis

Gene-based association statistics derived from exome sequencing data were obtained for schizophrenia (P-values in 'P meta' column from Table S5 from (*15*)), ASD (Q-values in 'qval_dnccPTV' column from Table S2 of (*34*)), and DD (P-values in 'denovoWEST_p_full' column from Table S2 of (*35*)). For each phenotype and each network, a one-tailed KS test was used to test whether the stringent interactors in the network have more significant association scores (i.e., smaller P-values or Q-values) compared to the non-interactors.

Transcriptional perturbation enrichment analysis

Differentially expressed genes (DEGs) between schizophrenia patients vs. controls identified in 20 prefrontal cortex cell types were retrieved from Supplementary Table 6 of (*12*).

The up- and down-regulated DEGs were analyzed as a joint set in the primary analysis, and then as separate sets in the secondary follow-up analyses. One-tailed P-values were calculated using a hypergeometric distribution to assess the overlap enrichment between interactors in our networks and the cell-type-specific DEGs. For each hypergeometric test, the 'population' was defined as all stringent interactor or non-interactor genes associated with a network and 'success in population' was defined as the stringent interactors in the network. The 'sample' contained DEGs in a cell type that were found in the population and 'success in sample' was the overlap between the interactors and the DEGs.

<u>Social Manhattan plot</u>

Genes mapped to 270 genome-wide significant regions in the combined discovery-replication meta-analysis of the PGC schizophrenia GWAS (phase 3) were retrieved from Supplementary Table 3 of the paper (*37*). We excluded non-coding genes and genes in the MHC region, and obtained the genomic positions of the remaining genes from Ensembl (*60*) (GRCh37.p13). We additionally included *SYNGAP1* by mapping it to the only MHC region SNP (rs13195636) included in the GWAS meta-analysis. Next, we intersected these GWAS genes with our interaction data to identify the subset of GWAS genes that are either an index gene or an interactor of an index gene. We then generated a Manhattan plot for these genes using the GWAS P-values of their associated index SNPs and plotted links between the genes to indicate protein-protein interactions in our data. In addition, we retrieved the list of GWAS genes prioritized by FINEMAP or SMR analysis from Supplementary Table 20 of the PGC paper and highlighted their overlap with genes prioritized by our data in the plot. For ease of visualization,

GWAS P-values were capped at $P$ = 1e-25 and genes in the PCDHA@ gene cluster were collapsed into one gene in the plot.

HCN1 network drug target query

To identify existing drugs targeting genes in the combined HCN1 network, we performed a batch query using the Open Targets Platform (*64*) (https://platform.opentargets.org). For each resulting drug-target entry, the corresponding drug mechanism information was retrieved from ChEMBL (*65*) (https://www.ebi.ac.uk/chembl/g/#browse/mechanisms_of_action).

**Text S1. Members of the Schizophrenia Working Group of the Psychiatric Genomics Consortium.**

Stephan Ripke [1,2] , Benjamin M Neale [1,2,3,4] , Aiden Corvin [5] , James TR Walters [6] , Kai-How Farh [1] , Peter A Holmans [6,7] , Phil Lee [1,2,4] , Brendan Bulik-Sullivan [1,2] , David A Collier [8,9] , Hailiang Huang [1,3] , Tune H Pers [3,10,11] , Ingrid Agartz [12,13,14] , Esben Agerbo [15,16,17] , Margot Albus [18] , Madeline Alexander [19] , Farooq Amin [20,21] , Silviu A Bacanu [22] , Martin Begemann [23] , Richard A Belliveau Jr [2] , Judit Bene [24,25] , Sarah E Bergen [2,26] , Elizabeth Bevilacqua [2] , Tim B Bigdeli [22] , Donald W Black [27] , Richard Bruggeman [28] , Nancy G Buccola [29] , Randy L Buckner [30,31,32] , William Byerley [33] , Wiepke Cahn [34] , Guiqing Cai [35,36] , Dominique Campion [37] , Rita M Cantor [38] , Vaughan J Carr [39,40] , Noa Carrera [6] , Stanley V Catts [39,41] , Kimberley D Chambert [2] , Raymond CK Chan [42] , Ronald YL Chan [43] , Eric YH Chen [44] , Wei Cheng [45] , Eric FC Cheung [46] , Siow Ann Chong [47] , C Robert Cloninger [48] , David Cohen [49] , Nadine Cohen [50] , Paul Cormican [5] , Nick Craddock [6,7] , James J Crowley [51] , David Curtis [52,53] , Michael Davidson [54] , Kenneth L Davis [36] , Franziska Degenhardt [55,56] , Jurgen Del Favero [57] , Ditte Demontis [17,58,59] , Dimitris Dikeos [60] , Timothy Dinan [61] , Srdjan Djurovic [14,62] , Gary Donohoe [5,63] , Elodie Drapeau [36] , Jubao Duan [64,65] , Frank Dudbridge [66] , Naser Durmishi [67] , Peter Eichhammer [68] , Johan Eriksson [69,70,71] , Valentina Escott-Price [6] , Laurent Essioux [72] , Ayman H Fanous [73,74,75,76] , Martilias S Farrell [51] , Josef Frank [77] , Lude Franke [78] , Robert Freedman [79] , Nelson B Freimer [80] , Marion Friedl [81] , Joseph I Friedman [36] , Menachem Fromer [1,2,4,82] , Giulio Genovese [2] , Lyudmila Georgieva [6] , Ina Giegling [81,83] , Paola Giusti-Rodríguez [51] , Stephanie Godard [84] , Jacqueline I Goldstein [1,3] , Vera Golimbet [85] , Srihari Gopal [86] , Jacob Gratten [87] , Lieuwe de Haan [88] , Christian Hammer [23] , Marian L Hamshere [6] , Mark Hansen [89] , Thomas Hansen [17,90] , Vahram Haroutunian [36,91,92] , Annette M Hartmann [81] , Frans A Henskens [39,93,94] , Stefan Herms [55,56,95] , Joel N Hirschhorn [3,11,96] , Per Hoffmann [55,56,95] , Andrea Hofman [55,56] , Mads V Hollegaard [97] , David M Hougaard [97] , Masashi Ikeda [98] , Inge Joa [99] , Antonio Julià [100] , René S Kahn [101] , Luba Kalaydjieva [102,103] , Sena Karachanak-Yankova [104] , Juha Karjalainen [78] , David Kavanagh [6] , Matthew C Keller [105] , James L Kennedy [106,107,108] , Andrey Khrunin [109] , Yunjung Kim [51] , Janis Klovins [110] , James A Knowles [111] , Bettina Konte [81] , Vaidutis Kucinskas [112] , Zita Ausrele Kucinskiene [112] , Hana Kuzelova-Ptackova [113,114] , Anna K Kähler [26] , Claudine Laurent [19,115] , Jimmy Lee [47,116] , S Hong Lee [87] , Sophie E Legge [6] , Bernard Lerer [117] , Miaoxin Li [118] , Tao Li [119] , Kung-Yee Liang [120] , Jeffrey Lieberman [121] , Svetlana Limborska [109] , Carmel M Loughland [39,122] , Jan Lubinski [123] , Jouko Lönnqvist [124] , Milan Macek [113,114] , Patrik KE Magnusson [26] , Brion S Maher [125] , Wolfgang Maier [126] , Jacques Mallet [127] , Sara Marsal [100] , Manuel Mattheisen [17,58,59,128] , Morten Mattingsdal [14,129] , Robert W McCarley [130,131] , Colm McDonald [132] , Andrew M McIntosh [133,134] , Sandra Meier [77] , Carin J Meijer [88] , Bela Melegh [24,25] , Ingrid Melle [14,135] , Raquelle I Mesholam-Gately [130,136] , Andres Metspalu [137] , Patricia T Michie [39,138] , Lili Milani [137] , Vihra Milanova [139] , Younes Mokrab [8] , Derek W Morris [5,63] , Ole Mors [17,58,140] , Kieran C Murphy [141] , Robin M Murray [142] , Inez Myin-Germeys [143] , Bertram Müller-Myhsok [144,145,146] , Mari Nelis [137] , Igor Nenadic [147] , Deborah A Nertney [148] , Gerald Nestadt [149] , Kristin K Nicodemus [150] , Liene Nikitina-Zake [110] , Laura Nisenbaum [151] , Annelie Nordin [152] , Eadbhard O'Callaghan [153] , Colm O'Dushlaine [2] , F Anthony O'Neill [154] , Sang-Yun Oh [155] , Ann Olincy [79] , Line Olsen [17,90] , Jim Van Os [143,156] , Psychosis Endophenotypes International Consortium [157] , Christos Pantelis [39,158] , George N Papadimitriou [60] , Sergi Papiol [23] , Elena Parkhomenko [36] , Michele T Pato [111] , Tiina Paunio [159,160] , Milica Pejovic-Milovancevic [161] , Diana O Perkins [162] , Olli Pietiläinen [160,163] , Jonathan Pimm [53] , Andrew J Pocklington [6] , John Powell [142] , Alkes Price

[164] , Ann E Pulver [149] , Shaun M Purcell [82] , Digby Quested [165] , Henrik B Rasmussen [17,90] , Abraham Reichenberg [36] , Mark A Reimers [166] , Alexander L Richards [6,7] , Joshua L Roffman [30,32] , Panos Roussos [82,167] , Douglas M Ruderfer [82] , Veikko Salomaa [71] , Alan R Sanders [64,65] , Ulrich Schall [39,122] , Christian R Schubert [168] , Thomas G Schulze [77,169] , Sibylle G Schwab [170] , Edward M Scolnick [2] , Rodney J Scott [39,171,172] , Larry J Seidman [130,136] , Jianxin Shi [173] , Engilbert Sigurdsson [174] , Teimuraz Silagadze [175] , Jeremy M Silverman [36,176] , Kang Sim [47] , Petr Slominsky [109] , Jordan W Smoller [2,4] , Hon-Cheong So [43] , Chris C A Spencer [177] , Eli A Stahl [3,82] , Hreinn Stefansson [178] , Stacy Steinberg [178] , Elisabeth Stogmann [179] , Richard E Straub [180] , Eric Strengman [181,182] , Jana Strohmaier [77] , T Scott Stroup [121] , Mythily Subramaniam [47] , Jaana Suvisaari [124] , Dragan M Svrakic [48] , Jin P Szatkiewicz [51] , Erik Söderman [12] , Srinivas Thirumalai [183] , Draga Toncheva [104] , Sarah Tosato [184] , Juha Veijola [185,186] , John Waddington [187] , Dermot Walsh [188] , Dai Wang [86] , Qiang Wang [119] , Bradley T Webb [22] , Mark Weiser [54] , Dieter B. Wildenauer [189] , Nigel M Williams [190] , Stephanie Williams [51] , Stephanie H Witt [77] , Aaron R Wolen [166] , Emily HM Wong [43] , Brandon K Wormley [22] , Hualin Simon Xi [191] , Clement C Zai [106,107] , Xuebin Zheng [192] , Fritz Zimprich [179] , Naomi R Wray [87] , Kari Stefansson [178] , Peter M Visscher [87] , Wellcome Trust Case-Control Consortium 2 [193] , Rolf Adolfsson [152] , Ole A Andreassen [14,135] , Douglas HR Blackwood [134] , Elvira Bramon [194] , Joseph D Buxbaum [35,36,91,195] , Anders D Børglum [17,58,59,140] , Sven Cichon [55,56,95,196] , Ariel Darvasi [197] , Enrico Domenici [198] , Hannelore Ehrenreich [23] , Tõnu Esko [3,11,96,137] , Pablo V Gejman [64,65] , Michael Gill [5] , Hugh Gurling [53] , Christina M Hultman [26] , Nakao Iwata [98] , Assen V Jablensky [39,199,200,201] , Erik G Jönsson [12] , Kenneth S Kendler [202] , George Kirov [6] , Jo Knight [106,107,108] , Todd Lencz [203,204,205] , Douglas F Levinson [19] , Qingqin S Li [86] , Jianjun Liu [192,206] , Anil K Malhotra [203,204,205] , Steven A McCarroll [2,96] , Andrew McQuillin [53] , Jennifer L Moran [2] , Preben B Mortensen [15,16,17] , Bryan J Mowry [87,207] , Markus M Nöthen [55,56] , Roel A Ophoff [38,80,208] , Michael J Owen [6,7] , Aarno Palotie [4,163,209] , Carlos N Pato [111] , Tracey L Petryshen [130,209,210] , Danielle Posthuma [211,212,213] , Marcella Rietschel [77] , Brien P Riley [202] , Dan Rujescu [81,83] , Pak C Sham [214] , Pamela Sklar [82,91,167] , David St Clair [215] , Daniel R Weinberger [180,216] , Jens R Wendland [168] , Thomas Werge [17,90,217] , Mark J Daly [1] , Patrick F Sullivan [26,51,162] , Michael C O'Donovan [6,7] .

[1] Analytic and Translational Genetics Unit, Massachusetts General Hospital, Boston, MA, USA. [2] Stanley Center for Psychiatric Research, Broad Institute of MIT and Harvard, Cambridge, MA, USA. [3] Medical and Population Genetics Program, Broad Institute of MIT and Harvard, Cambridge, MA, USA. [4] Psychiatric and Neurodevelopmental Genetics Unit, Massachusetts General Hospital, Boston, MA, USA. [5] Neuropsychiatric Genetics Research Group, Department of Psychiatry, Trinity College Dublin, Ireland. [6] MRC Centre for Neuropsychiatric Genetics and Genomics, Institute of Psychological Medicine and Clinical Neurosciences, School of Medicine, Cardiff University, Cardiff, UK. [7] National Centre for Mental Health, Cardiff University, Cardiff, Wales. [8] Eli Lilly and Company Limited, Erl Wood Manor, Sunninghill Road, Windlesham, Surrey, UK. [9] Social, Genetic and Developmental Psychiatry Centre, Institute of Psychiatry, King's College London, London, UK. [10] Center for Biological Sequence Analysis, Department of Systems Biology, Technical University of Denmark, Lyngby, Denmark. [11] Division of Endocrinology and Center for Basic and Translational Obesity Research, Boston Children's Hospital, Boston, MA, USA. [12] Department of Clinical Neuroscience, Karolinska Institutet, Stockholm, Sweden. [13] Department of Psychiatry, Diakonhjemmet Hospital, Oslo, Norway. [14] NORMENT, KG Jebsen Centre for Psychosis Research, Institute of Clinical Medicine, University of Oslo, Oslo, Norway. [15] Centre for Integrative Register-based Research, CIRRAU, Aarhus University, Aarhus, Denmark. [16] National Centre for Register-based Research, Aarhus

University, Aarhus, Denmark. [17] The Lundbeck Foundation Initiative for Integrative Psychiatric Research, iPSYCH, Denmark. [18] State Mental Hospital, Haar, Germany. [19] Department of Psychiatry and Behavioral Sciences, Stanford University, Stanford, CA, USA. [20] Department of Psychiatry and Behavioral Sciences, Atlanta Veterans Affairs Medical Center, Atlanta, GA, USA. [21] Department of Psychiatry and Behavioral Sciences, Emory University, Atlanta, GA, USA. [22] Virginia Institute for Psychiatric and Behavioral Genetics, Department of Psychiatry, Virginia Commonwealth University, Richmond, VA, USA. [23] Clinical Neuroscience, Max Planck Institute of Experimental Medicine, Göttingen, Germany. [24] Department of Medical Genetics, University of Pécs, Pécs, Hungary. [25] Szentagothai Research Center, University of Pécs, Pécs, Hungary. [26] Department of Medical Epidemiology and Biostatistics, Karolinska Institutet, Stockholm, Sweden. [27] Department of Psychiatry, University of Iowa Carver College of Medicine, Iowa City, IA, USA. [28] University Medical Center Groningen, Department of Psychiatry, University of Groningen, The Netherlands. [29] School of Nursing, Louisiana State University Health Sciences Center, New Orleans, LA, USA. [30] Athinoula A. Martinos Center, Massachusetts General Hospital, Boston, MA, USA. [31] Center for Brain Science, Harvard University, Cambridge MA, USA. [32] Department of Psychiatry, Massachusetts General Hospital, Boston, MA, USA. [33] Department of Psychiatry, University of California at San Francisco, San Francisco, CA, USA. [34] University Medical Center Utrecht, Department of Psychiatry, Rudolf Magnus Institute of Neuroscience, The Netherlands. [35] Department of Human Genetics, Icahn School of Medicine at Mount Sinai, New York, NY, USA. [36] Department of Psychiatry, Icahn School of Medicine at Mount Sinai, New York, NY, USA. [37] Centre Hospitalier du Rouvray and INSERM U1079 Faculty of Medicine, Rouen, France. [38] Department of Human Genetics, David Geffen School of Medicine, University of California, Los Angeles, CA, USA. [39] Schizophrenia Research Institute, Sydney, Australia. [40] School of Psychiatry, University of New South Wales, Sydney, Australia. [41] Royal Brisbane and Women's Hospital, University of Queensland, Brisbane, Australia. [42] Institute of Psychology, Chinese Academy of Science, Beijing, PR China. [43] Department of Psychiatry, Li Ka Shing Faculty of Medicine, The University of Hong Kong, Hong Kong SAR, PR China. [44] Department of Psychiatry and State Ket Laboratory for Brain and Cognitive Sciences, Li Ka Shing Faculty of Medicine, The University of Hong Kong, Hong Kong SAR, PR China. [45] Department of Computer Science, University of North Carolina, Chapel Hill, NC, USA. [46] Castle Peak Hospital, Hong Kong SAR, PR China. [47] Institute of Mental Health, Singapore. [48] Department of Psychiatry, Washington University, St. Louis, MO, USA. [49] Department of Child and Adolescent Psychiatry, Pierre and Marie Curie Faculty of Medicine and Brain and Spinal Cord Institute (ICM), Paris, France. [50] Formerly of Neuroscience Therapeutic Area, Janssen Research and Development, LLC, Raritan, NJ, USA. [51] Department of Genetics, University of North Carolina, Chapel Hill, NC, USA. [52] Department of Psychological Medicine, Queen Mary University of London, UK. [53] Molecular Psychiatry Laboratory, Division of Psychiatry, University College London, UK. [54] Sheba Medical Center, Tel Hashomer, Israel. [55] Department of Genomics, Life and Brain Center, Bonn, Germany. [56] Institute of Human Genetics, University of Bonn, Bonn, Germany. [57] Applied Molecular Genomics Unit, VIB Department of Molecular Genetics, University of Antwerp, Antwerp, Belgium. [58] Centre for Integrative Sequencing, iSEQ, Aarhus University, Aarhus, Denmark. [59] Department of Biomedicine, Aarhus University, Aarhus, Denmark. [60] First Department of Psychiatry, University of Athens Medical School, Athens, Greece. [61] Department of Psychiatry, University College Cork, Ireland. [62] Department of Medical Genetics, Oslo University Hospital, Oslo, Norway. [63] Cognitive Genetics and Therapy Group, School of Psychology and Discipline of Biochemistry,

National University of Ireland Galway, Ireland. [64] Department of Psychiatry and Behavioral Neuroscience, University of Chicago, Chicago, IL, USA. [65] Department of Psychiatry and Behavioral Sciences, NorthShore University HealthSystem, Evanston, IL, USA. [66] Department of Non-Communicable Disease Epidemiology, London School of Hygiene and Tropical Medicine, London, UK. [67] Department of Child and Adolescent Psychiatry, University Clinic of Psychiatry, Skopje, Republic of Macedonia. [68] Department of Psychiatry, University of Regensburg, Regensburg, Germany. [69] Department of General Practice, Helsinki University Central Hospital, Helsinki, Finland. [70] Folkhälsan Research Center, Helsinki, Finland. [71] National Institute for Health and Welfare, Helsinki, Finland. [72] Translational Technologies and Bioinformatics, Pharma Research and Early Development, F.Hoffman-La Roche, Basel, Switzerland. [73] Department of Psychiatry, Georgetown University School of Medicine, Washington DC, USA. [74] Department of Psychiatry, Keck School of Medicine of the University of Southern California, Los Angeles, CA, USA. [75] Department of Psychiatry, Virginia Commonwealth University School of Medicine, Richmond, VA, USA. [76] Mental Health Service Line, Washington VA Medical Center, Washington DC, USA. [77] Department of Genetic Epidemiology in Psychiatry, Central Institute of Mental Health, Medical Faculty Mannheim, University of Heidelberg, Heidelberg, Germany. [78] Department of Genetics, University of Groningen, University Medical Centre Groningen, The Netherlands. [79] Department of Psychiatry, University of Colorado Denver, Aurora, CO, USA. [80] Center for Neurobehavioral Genetics, Semel Institute for Neuroscience and Human Behavior, University of California, Los Angeles, CA, USA. [81] Department of Psychiatry, University of Halle, Halle, Germany. [82] Division of Psychiatric Genomics, Department of Psychiatry, Icahn School of Medicine at Mount Sinai, New York, NY, USA. [83] Department of Psychiatry, University of Munich, Munich, Germany. [84] Departments of Psychiatry and Human and Molecular Genetics, INSERM, Institut de Myologie, Hôpital de la Pitiè-Salpêtrière, Paris, France. [85] Mental Health Research Centre, Russian Academy of Medical Sciences, Moscow, Russia. [86] Neuroscience Therapeutic Area, Janssen Research and Development, LLC, Raritan, NJ, USA. [87] Queensland Brain Institute, The University of Queensland, Brisbane, Queensland, Australia. [88] Academic Medical Centre University of Amsterdam, Department of Psychiatry, Amsterdam, The Netherlands. [89] Illumina, Inc., La Jolla, CA, USA. [90] Institute of Biological Psychiatry, MHC Sct. Hans, Mental Health Services Copenhagen, Denmark. [91] Friedman Brain Institute, Icahn School of Medicine at Mount Sinai, New York, NY, USA. [92] JJ Peters VA Medical Center, Bronx, NY, USA. [93] Priority Research Centre for Health Behaviour, University of Newcastle, Newcastle, Australia. [94] School of Electrical Engineering and Computer Science, University of Newcastle, Newcastle, Australia. [95] Division of Medical Genetics, Department of Biomedicine, University of Basel, Basel, Switzerland. [96] Department of Genetics, Harvard Medical School, Boston, MA, USA. [97] Section of Neonatal Screening and Hormones, Department of Clinical Biochemistry, Immunology and Genetics, Statens Serum Institut, Copenhagen, Denmark. [98] Department of Psychiatry, Fujita Health University School of Medicine, Toyoake, Aichi, Japan. [99] Regional Centre for Clinical Research in Psychosis, Department of Psychiatry, Stavanger University Hospital, Stavanger, Norway. [100] Rheumatology Research Group, Vall d'Hebron Research Institute, Barcelona, Spain. [101] Department of Psychiatry, Rudolf Magnus Institute of Neuroscience, University Medical Center Utrecht, Utrecht, The Netherlands. [102] Centre for Medical Research, The University of Western Australia, Perth, Western Australia, Australia. [103] Perkins Institute for Medical Research, The University of Western Australia, Perth, Western Australia, Australia. [104] Department of Medical Genetics, Medical University, Sofia, Bulgaria. [105] Department of Psychology, University of Colorado

Boulder, Boulder, CO, USA. [106] Campbell Family Mental Health Research Institute, Centre for Addiction and Mental Health, Toronto, Ontario, Canada. [107] Department of Psychiatry, University of Toronto, Toronto, Ontario, Canada. [108] Institute of Medical Science, University of Toronto, Toronto, Ontario, Canada. [109] Institute of Molecular Genetics, Russian Academy of Sciences, Moscow, Russia. [110] Latvian Biomedical Research and Study Centre, Riga, Latvia. [111] Department of Psychiatry and Zilkha Neurogenetics Institute, Keck School of Medicine at University of Southern California, Los Angeles, CA, USA. [112] Faculty of Medicine, Vilnius University, Vilnius, Lithuania. [113] 2nd Faculty of Medicine and University Hospital Motol, Prague, Czech Republic. [114] Department of Biology and Medical Genetics, Charles University Prague, Prague, Czech Republic. [115] Pierre and Marie Curie Faculty of Medicine, Paris, France. [116] Duke-NUS Graduate Medical School, Singapore. [117] Department of Psychiatry, Hadassah-Hebrew University Medical Center, Jerusalem, Israel. [118] Centre for Genomic Sciences and Department of Psychiatry, Li Ka Shing Faculty of Medicine, The University of Hong Kong, Hong Kong SAR, PR China. [119] Mental Health Centre and Psychiatric Laboratory, West China Hospital, Sichuan University, Chendu, Sichuan, PR China. [120] Department of Biostatistics, Johns Hopkins University Bloomberg School of Public Health, Baltimore, Maryland, USA. [121] Department of Psychiatry, Columbia University, New York, NY, USA. [122] Priority Centre for Translational Neuroscience and Mental Health, University of Newcastle, Newcastle, Australia. [123] Department of Genetics and Pathology, International Hereditary Cancer Center, Pomeranian Medical University in Szczecin, Szczecin, Poland. [124] Department of Mental Health and Substance Abuse Services; National Institute for Health and Welfare, Helsinki, Finland. [125] Department of Mental Health, Bloomberg School of Public Health, Johns Hopkins University, Baltimore, MD, USA. [126] Department of Psychiatry, University of Bonn, Bonn, Germany. [127] Centre National de la Recherche Scientifique, Laboratoire de Génétique Moléculaire de la Neurotransmission et des Processus Neurodégénératifs, Hôpital de la Pitié Salpêtrière, Paris, France. [128] Department of Genomics Mathematics, University of Bonn, Bonn, Germany. [129] Research Unit, Sørlandet Hospital, Kristiansand, Norway. [130] Department of Psychiatry, Harvard Medical School, Boston, MA, USA. [131] VA Boston Health Care System, Brockton, MA, USA. [132] Department of Psychiatry, National University of Ireland Galway, Ireland. [133] Centre for Cognitive Ageing and Cognitive Epidemiology, University of Edinburgh, UK. [134] Division of Psychiatry, University of Edinburgh, Edinburgh, UK. [135] Division of Mental Health and Addiction, Oslo University Hospital, Oslo, Norway. [136] Massachusetts Mental Health Center Public Psychiatry Division of the Beth Israel Deaconess Medical Center, Boston, MA, USA. [137] Estonian Genome Center, University of Tartu, Tartu, Estonia. [138] School of Psychology, University of Newcastle, Newcastle, Australia. [139] First Psychiatric Clinic, Medical University, Sofia, Bulgaria. [140] Department P, Aarhus University Hospital, Risskov, Denmark. [141] Department of Psychiatry, Royal College of Surgeons in Ireland, Ireland. [142] King's College London, UK. [143] Maastricht University Medical Centre, South Limburg Mental Health Research and Teaching Network, EURON, Maastricht, The Netherlands. [144] Institute of Translational Medicine, University Liverpool, UK. [145] Max Planck Institute of Psychiatry, Munich, Germany. [146] Munich Cluster for Systems Neurology (SyNergy), Munich, Germany. [147] Department of Psychiatry and Psychotherapy, Jena University Hospital, Jena, Germany. [148] Department of Psychiatry, Queensland Brain Institute and Queensland Centre for Mental Health Research, University of Queensland, Brisbane, Queensland, Australia. [149] Department of Psychiatry and Behavioral Sciences, Johns Hopkins University School of Medicine, Baltimore, Maryland, USA. [150] Department of Psychiatry, Trinity College Dublin, Ireland. [151] Eli Lilly and Company, Lilly

Corporate Center, Indianapolis, IN, USA. [152] Department of Clinical Sciences, Psychiatry, Umeå University, Umeå, Sweden. [153] DETECT Early Intervention Service for Psychosis, Blackrock, Dublin, Ireland. [154] Centre for Public Health, Institute of Clinical Sciences, Queens University Belfast, Belfast, UK. [155] Lawrence Berkeley National Laboratory, University of California at Berkeley, Berkeley, CA, USA. [156] Institute of Psychiatry at King's College London, London, UK. [157] PEIC. [158] Melbourne Neuropsychiatry Centre, University of Melbourne & Melbourne Health, Melbourne, Australia. [159] Department of Psychiatry, University of Helsinki, Finland. [160] Public Health Genomics Unit, National Institute for Health and Welfare, Helsinki, Finland. [161] Medical Faculty, University of Belgrade, Belgrade, Serbia. [162] Department of Psychiatry, University of North Carolina, Chapel Hill, NC, USA. [163] Institute for Molecular Medicine Finland, FIMM, Helsinki, Finland. [164] Department of Epidemiology, Harvard University, Boston, MA, USA. [165] Department of Psychiatry, University of Oxford, Oxford, UK. [166] Virginia Institute for Psychiatric and Behavioral Genetics, Virginia Commonwealth University, Richmond, VA, USA. [167] Institute for Multiscale Biology, Icahn School of Medicine at Mount Sinai, New York, NY, USA. [168] PharmaTherapeutics Clinical Research, Pfizer Worldwide Research and Development, Cambridge, MA, USA. [169] Department of Psychiatry and Psychotherapy, University of Gottingen, Göttingen, Germany. [170] Psychiatry and Psychotherapy Clinic, University of Erlangen, Germany. [171] Hunter New England Health Service, Newcastle, Australia. [172] School of Biomedical Sciences, University of Newcastle, Newcastle, Australia. [173] Division of Cancer Epidemiology and Genetics, National Cancer Institute, Bethesda, MD, USA. [174] University of Iceland, Landspitali, National University Hospital, Reykjavik, Iceland. [175] Department of Psychiatry and Drug Addiction, Tbilisi State Medical University (TSMU), Tbilisi, Georgia. [176] Research and Development, Bronx Veterans Affairs Medical Center, New York, NY, USA. [177] Wellcome Trust Centre for Human Genetics, Oxford, UK. [178] deCODE Genetics, Reykjavik, Iceland. [179] Department of Clinical Neurology, Medical University of Vienna, Austria. [180] Lieber Institute for Brain Development, Baltimore, MD, USA. [181] Department of Medical Genetics, University Medical Centre, Utrecht, The Netherlands. [182] Rudolf Magnus Institute of Neuroscience, University Medical Centre Utrecht, The Netherlands. [183] Berkshire Healthcare NHS Foundation Trust, Bracknell, UK. [184] Section of Psychiatry, University of Verona, Verona, Italy. [185] Department of Psychiatry, University of Oulu, Finland. [186] University Hospital of Oulu, Oulu, Finland. [187] Molecular and Cellular Therapeutics, Royal College of Surgeons in Ireland, Dublin, Ireland. [188] Health Research Board, Dublin, Ireland. [189] Department of Psychiatry and Clinical Neurosciences, School of Psychiatry and Clinical Neurosciences, Queen Elizabeth II Medical Centre, Perth, Western Australia, Australia. [190] Department of Psychological Medicine and Neurology, MRC Centre for Neuropsychiatric Genetics and Genomics, School of Medicine, Cardiff University, Cardiff, Wales, UK. [191] Computational Sciences CoE, Pfizer Worldwide Research and Development, Cambridge, MA, USA. [192] Human Genetics, Genome Institute of Singapore, A*STAR, Singapore. [193] WTCCC2. [194] University College London, UK. [195] Department of Neuroscience, Icahn School of Medicine at Mount Sinai, New York, NY, USA. [196] Institute of Neuroscience and Medicine (INM-1), Research Center Juelich, Juelich, Germany. [197] Department of Genetics, The Hebrew University of Jerusalem, Jerusalem, Israel. [198] Neuroscience Discovery and Translational Area, Pharma Research and Early Development, F.Hoffman-La Roche, Basel, Switzerland. [199] School of Psychiatry and Clinical Neurosciences, The University of Western Australia, Perth, Australia. [200] The Perkins Institute of Medical Research, Perth, Australia. [201] UWA Centre for Clinical Research in Neuropsychiatry. [202] Virginia Institute for Psychiatric and Behavioral Genetics, Departments of Psychiatry and

Human and Molecular Genetics, Virginia Commonwealth University, Richmond, VA, USA. [203] The Feinstein Institute for Medical Research, Manhasset, NY, USA. [204] The Hofstra NS-LIJ School of Medicine, Hempstead, NY, USA. [205] The Zucker Hillside Hospital, Glen Oaks, NY, USA. [206] Saw Swee Hock School of Public Health, National University of Singapore, Singapore. [207] Queensland Centre for Mental Health Research, University of Queensland, Brisbane, Queensland, Australia. [208] Department of Psychiatry, Brain Center Rudolf Magnus, University Medical Center Utrecht, The Netherlands. [209] The Broad Institute of MIT and Harvard, Cambridge, MA, USA. [210] Center for Human Genetic Research and Department of Psychiatry, Massachusetts General Hospital, Boston, MA, USA. [211] Department of Child and Adolescent Psychiatry, Erasmus University Medical Centre, Rotterdam, The Netherlands. [212] Department of Complex Trait Genetics, Neuroscience Campus Amsterdam, VU University Medical Center Amsterdam, Amsterdam, The Netherlands. [213] Department of Functional Genomics, Center for Neurogenomics and Cognitive Research, Neuroscience Campus Amsterdam, VU University, Amsterdam, The Netherlands. [214] Centre for Genomic Sciences, State Ket Laboratory for Brain and Cognitive Sciences, and Department of Psychiatry, Li Ka Shing Faculty of Medicine, The University of Hong Kong, Hong Kong SAR, PR China. [215] University of Aberdeen, Institute of Medical Sciences, Aberdeen, Scotland, UK. [216] Departments of Psychiatry, Neurology, Neuroscience and Institute of Genetic Medicine, Johns Hopkins School of Medicine, Baltimore, MD, USA. [217] Department of Clinical Medicine, University of Copenhagen, Copenhagen, Denmark.

**Text S2. Members of the Stanley Center Asia Initiatives.**

| Name | Institution |
|---|---|
| Hailiang Huang | Broad Institute of MIT and Harvard, USA |
| Shengying Qin | Shanghai Jiao Tong University Bio-X Institute, China |
| Akira Sawa | The Johns Hopkins University School of Medicine and Bloomberg School of Public Health, the Johns Hopkins Hospital, USA |
| Sibylle G Schwab | School of Chemistry and Molecular Bioscience, University of Wollongong<br>Illawarra Health and Medical Research Institute, Wollongong, Australia |
| Rene Kahn | Icahn School of Medicine at Mount Sinai, USA |
| Kyung Sue Hong | Department of Psychiatry, Sungkyunkwan University, Samsung Medical Center, Seoul, Korea |
| Wenzhao Shi | Digital China Health, China |
| Ming Tsuang | University of California San Diego, USA |
| Masanari Itokawa | Tokyo Metropolitan Institute of Medical Science, Japan |
| Gang Feng | Digital China Health, China |
| Jianjun Liu | Genome Institute of Singapore, A*STAR<br>Yong Loo Lin School of Medicine, National University of Singapore, Singapore |
| Stephen J. Glatt | SUNY Upstate Medical University, USA |
| Nakao Iwata | Fujita Health University School of Medicine, Japan |
| Masashi Ikeda | Fujita Health University School of Medicine, Japan |
| Xiancang Ma | Xi'an Jiao Tong University, China |
| Jimmy Lee | Institute of Mental Health, Singapore,<br>Lee Kong Chian School of Medicine, Nanyang Technological University, Singapore |
| Jinsong Tang | Zhejiang University, China |
| Yunfeng Ruan | Broad Institute of MIT and Harvard, USA |
| Ruize Liu | Broad Institute of MIT and Harvard, USA |
| Feng Zhu | Xi'an Jiao Tong University, China |
| Yasue Horiuchi | Tokyo Metropolitan Institute of Medical Science, Tokyo, Japan |
| Byung Dae Lee | Department of Psychiatry, Pusan National University Hospital, Korea |
| Eun-Jeong Joo | Department of Neuropsychiatry School of Medicine, Eulji University, Korea |

| | |
|---|---|
| Woojae Myung | Department of Psychiatry, Seoul National University Bundang Hospital, Korea |
| Kyooseob Ha | Department of Psychiatry, Seoul National University Hospital, Korea |
| Hong-Hee Won | Sungkyunkwan University, Korea |
| Ji Hyung Baek | Department of Psychiatry, Sungkyunkwan University, Samsung Medical Center, Korea |
| Young Chul Chung | Department of Psychiatry, Chonbuk National University Medical School, Korea |
| Sung-Wan Kim | Department of Psychiatry, Chonnam National University Medical School, Korea |
| Dieter B Wildenauer | University of Western Australia, Perth, Australia |
| Agung Kusumawardhani | Department of Psychiatry, University of Indonesia, Jakarta, Indonesia |
| Wei J. Chen | National Taiwan U. |
| Hai-Gwo Hwu | National Taiwan U. |
| Kang Sim | Institute of Mental Health, Singapore |
| Akitoyo Hishimoto | Kobe University Graduate School of Medicine, Japan |
| Ikuo Otsuka | Kobe University Graduate School of Medicine, Japan |
| Ichiro Sora | Kobe University Graduate School of Medicine, Japan |
| Tomoko Toyota | RIKEN Center for Brain Science, Wako, Japan |
| Takeo Yoshikawa | RIKEN Center for Brain Science, Wako, Japan |
| Hiroshi Kunugi | National Institute of Neuroscience, National Center of Neurology and Psychiatry, Japan. |
| Kotaro Hattori | National Institute of Neuroscience, National Center of Neurology and Psychiatry, Japan. |
| Sayuri Ishiwata | National Institute of Neuroscience, National Center of Neurology and Psychiatry, Japan. |
| Shusuke Numata | Department of Psychiatry, Institute of Biomedical Sciences, Tokushima University Graduate School, Tokushima, Japan |
| Tetsuro Ohmori | Department of Psychiatry, Institute of Biomedical Sciences, Tokushima University Graduate School, Tokushima, Japan |
| Makoto Arai | Tokyo Metropolitan Institute of Medical Science, Tokyo, Japan |
| Yuji Ozeki | Department of Psychiatry, Shiga University of Medical Science, Japan |
| Kumiko Fujii | Department of Psychiatry, Shiga University of Medical Science, Japan |
| Se Joo Kim | Department of Psychiatry, Yonsei University College of Medicine, |

| | |
|---|---|
| | Korea |
| Heon-Jeong Lee | Department of Psychiatry, Korea University College of Medicine, Korea |
| Yong Min Ahn | Department of Psychiatry, Seoul National University Hospital, Korea |
| Se Hyun Kim | Department of Psychiatry, Seoul National University Hospital, Korea |
| Kazufumi Akiyama | Department of Biological Psychiatry and Neuroscience, Dokkyo Medical University, School of Medicine, Japan |
| Kazutaka Shimoda | Department of Psychiatry, Dokkyo Medical University School of Medicine, Japan |
| Makoto Kinoshita | Department of Psychiatry, Institute of Biomedical Sciences, Tokushima University Graduate School, Tokushima, Japan |

**Text S3. IRBs or oversight bodies that provided approval or exemption for human subjects data used in this study.**

IRB approvals and study consent forms from each of the sample contributing organizations were sent to the Broad Institute before samples were sequenced and analyzed. All ethical approvals are on file at the Massachusetts General Brigham (MGB), formerly Partners, IRB office amended to protocol #2014P001342, title: 'Molecular Profiling of Psychiatric Disease'. UK Biobank (UKBB) GWAS summary statistics were released by the Neale Lab and are publicly available (http://www.nealelab.is/uk-biobank). Biobank Japan (BBJ) GWAS summary statistics are publicly available on the BBJ website (http://jenger.riken.jp/en/result). Schizophrenia, ADHD, ASD, BIP, and MDD GWAS summary statistics are publicly available on the Psychiatric Genomics Consortium (PGC) website (https://www.med.unc.edu/pgc/download-results/). The use of schizophrenia individual-level data of European and East Asian ancestry in the present work were approved by the PGC and the Stanley Global Asia Initiatives, respectively. The following institutions provided ethics oversight for the collection of the data: regional Ethical Review Board at University of Umeå; Norwegian Scientific-Ethical Committee and the Norwegian Data Protection Agency; Scotland A Research Ethics Committee; Danish Data Protection Agency and the ethics committees of Denmark; Psychosis Endophenotypes International Consortium (PEIC); King's College London; University of Amsterdam; University of Groningen; Maastricht University Medical Centre; University of Utrecht; Mount Sinai; Federated Dublin Hospitals and Irish Blood Transfusion Services Research Ethics Committees; Irish Blood Transfusion Service; Georg-August-University; University of Tartu; Molecular Genetics of Schizophrenia (MGS); University College London and UK National Health Service multicenter; Karolinska Hospital and the Stockholm Regional Ethical Committee; Karolinska Institute; Bulgaria ethics committees; Hebrew University Genetic Resource and National Genetic Committee of the Israeli Ministry of Health; Wellcome Trust Case Control Consortium; North Shore-Long Island Jewish Health System; Australian Schizophrenia Research Bank; Multicentre Research Ethics Committee in Wales; UK Multicentre Research Ethics Committee (MREC); University Medical Centre Utrecht; SUNY Upstate Medical University; MooDS Consortium; Clinical Antipsychotics Trials of Intervention Effectiveness (CATIE); Clinical Brain Disorders Branch of the NIMH; Domain Specific Review Board (Singapore); Fujita Health University; RIKEN Center for Integrative Medical Sciences; Nagoya University; Osaka University; Niigata University; University Medical Center Utrecht; The University of Western Australia; The University of Indonesia; National Taiwan University; Samsung Medical Center; Chonnam National University Hospital; Tokyo Metropolitan Institute of Medical Science and affiliated institutions; Harvard T.H. Chan School of Public Health; Partners Institutional Review Boards.

**Text S4. Prenatal vs. postnatal expression of schizophrenia risk genes.**

To further explore the pre- vs. postnatal expression differences of schizophrenia risk genes identified in Set 3 compared to the SCHEMA genes in **Fig. 1C**, we first referred to the SCHEMA preprint (*15*), in which Singh and colleagues explored what properties may differ between schizophrenia and severe neurodevelopmental disorders (DD/ID) risk genes implicated by rare variants. Remarkably, they found that the DD/ID genes are under stronger evolutionary constraint and have a bias towards prenatal expression. Furthermore, they showed that even though schizophrenia is a later-onset disorder, risk genes that are impacted by ultra rare loss-of-function mutations in this disease significantly overlap with DD/ID and do not show pre- or postnatal expression bias as a group (i.e., schizophrenia genes are evenly distributed when ranked by degree of pre- to postnatal expression against all genes; Fig. S23 of the preprint).

Building on these genetic observations, a possible explanation for the lower prenatal expression of our Set 3 genes compared to the SCHEMA genes is that schizophrenia genes are influenced by risk variants across the entire spectrum of allele frequencies (from ultra rare to very common) and that our index gene selection approach from GWAS data likely lead to a set of schizophrenia genes more on the ‘common variant’ spectrum. This means that they are likely less constrained than the SCHEMA genes and are perhaps less critical during neurodevelopment, leading to lower prenatal expression as observed in **Fig. 1C**. This is speculation, however, and we did not observe a significant difference between Set 3 vs. the exome-wide significant SCHEMA genes when we compared their gnomAD pLI scores (two-tailed KS test $P = 0.23$) and their DD/ID association scores (two-tailed KS test $P = 0.83$). Regardless, it is still a plausible hypothesis that the SCHEMA genes prioritized from rare variants with large effect sizes may capture more of the neurodevelopmental aspects of schizophrenia-related biology compared to the Set 3 genes implicated by common variants, which perhaps reflect later processes in neuronal maturation and plasticity during adolescence.

To further investigate this issue, we compared the individual BrainSpan expression of the top SCHEMA genes vs. our six index genes included in the final PPI network. On a single-gene level, we did not see a consistent pre- or postnatal bias trend in either group of genes (**Fig. S7A**). Among the SCHEMA genes, *CACNA1G* and *GRIN2A*, which have also been associated with DD/ID, actually show strong postnatal bias, while the other genes have consistently high prenatal and postnatal expression. Among the index genes, none have a very pronounced pattern, even though a few genes have slightly higher expression postnatally. Conversely, several of the Set 3 genes that originally contributed to the strong postnatal bias observed in **Fig. 1C** were excluded from the PPI network due to a lack of IP-competent antibodies. Therefore, the expression profiles of the six index genes that we included in the network generally agree with the expression profiles of all network genes, which show no obvious pre- or postnatal bias and are similar to the SCHEMA genes (**Fig. 4C** and **Fig. S7A**).

In summary, for all different schizophrenia gene sets that we have analyzed, there is not one consistent pre- and postnatal expression profile, as can be expected for a complex disorder like schizophrenia, and the differences in expression trends that we observed when grouping each set of genes and comparing at a group level is an oversimplification of the underlying expression profiles at the level of individual genes.

**Text S5. Discussion on the $\log_2$ FC cutoff used to define index protein interactors.**

In proteomic data, the distribution of observed fold changes (FCs) is directly dependent on the experimental design in a given study. That is, FC is a relative, context-dependent measure and often not directly comparable between studies. For example, as discussed in **Text S6**, many previous interaction proteomics studies have aimed at identifying core molecular complexes through tagging and overexpressing proteins for immunoprecipitation, mostly in 'generic' cell types that are easy to transfect. In these studies, it may be more common to detect interactions at high FCs (>2) due to overexpression of proteins (*66*). In contrast, we purposely designed our IP experiments to pull down endogenously expressed protein isoforms in neurons, some of which may have relatively low expression despite their functional importance (e.g., SYNGAP1 (*67*)); we also titrated our IP conditions to preserve larger and more inclusive networks of interacting proteins rather than restricting to narrowly defined core protein complexes that capture more limited biological insights. These design choices directly impact the measured protein abundances and FCs observed in our experiments and may result in relatively low FCs. In addition, differences in MS methods would also be a major factor determining the FC observed in the experiment. For example, data generated using isobaric labeling tend to have lower FCs due to ratio compression compared to those generated with a label-free approach. This is indeed what we observed in the accompanying ASD study, where interactors identified using isobaric labeling MS have a median $\log_2$ FC of ~1, compared to a median of ~5 for those from label-free data.

Therefore, we chose not to use an arbitrary FC cutoff when defining significant interactions as it would not be consistent across studies and protocols. Instead, we defined all proteins with $\log_2$ FC > 0 and FDR $\leq$ 0.1 as significant interactors of the index protein in each IP-MS experiment. However, we note that the statistical method that we used to calculate significance (i.e., the moderated t-test implemented in the Genoppi/limma R package) is inherently designed so that proteins with larger FC (and thus larger sample variance) are more likely to be significant compared to proteins with smaller FC (*68*). Similar concepts were implemented in several other established approaches for mapping PPIs that implicitly consider FC information, but do not specifically use an FC cutoff (e.g., ComPASS-Plus used in BioPlex (*69*)).

In addition, we performed validation experiments and computational analyses to support the high quality and biological relevance of the identified interactors regardless of their FCs:

1. **Reverse IP experiments validated interactions across a wide range of FCs.** To assess the quality and reproducibility of our PPI data, we have performed a significant amount of validation experiments including both forward and reverse IPs, testing interactions identified with $\log_2$ FCs of 0.12 to 2.23 (**Fig. 2C**, **Fig. S3-5**, and **Data S7**). In particular, we performed reverse IPs using a panel of interactors as baits, and successfully detected the original index proteins in 23 out of 25 reverse IPs that showed bait enrichment (92.0% validation rate). Given that we do not expect all true interactions to be detectable bidirectionally, these results support a low false positive rate (~8%) in our data that is in remarkable agreement with the 10% FDR cutoff we applied to identify significant interactions. We also specifically tested whether there was any association between FCs in the original IP-MS data and validation outcomes in the reverse IPs and did not observe any significant relationship; in fact, we were able to validate interactors identified at $\log_2$ FCs of 0.12 to 1.85 (**Data S7**).

2. **Additional validation using known protein interaction data.** We found that interactors in 10 of the 19 IPs are enriched for known InWeb interactors, including some gold standard interactors that have relatively low FCs in our data (**Data S5-6**). For example, in the CACNA1C IP highlighted in **Fig. 2A**,**B**, all of the known L-type calcium channel subunits that form a complex with the transmembrane CACNA1C were identified as significant interactors, with $\log_2$ FC of 0.7 for the extracellular CACNA2D1 and $\log_2$ FCs of 1.5 and 1.6 for the intracellular CACNB1 and CACNB3, respectively. The relatively low FCs here are certainly not reflective of these interactors being false positives as they are clearly biologically valid. Rather they reflect our experimental design and the isobaric labeling MS protocol used to generate the data.

3. **Orthogonal support using brain co-expression data.** We used human and mouse brain co-expression data to show that on average, transcripts of the interactors are more likely to be co-expressed (using bulk or single-cell RNA-seq data) or co-localized (using spatial transcriptomic data) with that of the index proteins compared to i) the non-interactors [i.e., non-significant proteins with $\log_2$ FC $\leq 0$ or FDR $> 0.1$ in IP-MS]; ii) known interactors in InWeb; and iii) all protein-coding genes (**Fig. 2D** and **Fig. S2C**). Another key observation is that when we grouped interactors based on different $\log_2$ FC thresholds, we did not observe a positive association between their FCs and co-expression scores in three out of the four tested expression datasets (**Fig. S2D**). This suggests that the interactions identified at lower FCs could have as much biological relevance as those identified at higher FCs in terms of representing pairwise functional gene relationships in complex brain tissues.

In conclusion, FCs are strongly dependent on the experimental design and our deliberate choices in this study would not result in very high FC values. We also note that, since FCs are context-dependent, there is no consensus in the literature about what a relevant or appropriate global FC cutoff would be in IP-MS experiments. However, the experimental and computational validations that we have described above strongly support the high quality of our data despite using an inclusive FC cutoff to define interactors across our experiments. Furthermore, our results indicate that there are no obvious quality differences between the interactors we identified across a range of different FCs.

**Text S6. Factors contributing to the large percentage of novel interactions in our data.**

Across the 19 IP-MS datasets we analyzed, > 96% of the protein interactions have not been reported in InWeb. The large percentage of novel interactions can readily be explained by a combination of the brain cell-type-specificity of the data we report here and several additional deliberate design choices in our experimental protocol (which substantially differs from some common experimental approaches underlying data in InWeb). The design choices in question include: i) immunoprecipitation of endogenous [i.e., non-tagged and non-overexpressed] protein isoforms expressed in neurons; ii) IPs performed in a salt/detergent environment optimized for identifying inclusive protein networks instead of narrowly defined core protein complexes; and iii) IPs performed exclusively in neurons and not other 'generic' cell types. In more detail:

1. We performed IPs of endogenously expressed proteins using commercially available immunoreagents to avoid artifacts associated with tagged overexpression systems (e.g., due to excess of tagged proteins, biochemical interference of the tag with the interactions of the bait protein, or arbitrary selection of a specific protein isoform for tagging and overexpressing). This design choice allowed us to study relevant proteins in neurons, which are difficult-to-transfect cell types, without biases for previously annotated isoforms. Additionally, we did not perform any cross-linking prior to immunoprecipitation, which usually drives enrichment of the 'bait' protein.

2. Our experimental strategy was also designed to identify inclusive protein networks not limited to stringently defined core protein complexes. This means that we are deliberately including both first- and second-order interactions in our data. We achieved this by titrating detergent composition during co-IP washes (NP-40 is 0.01% v/v) and by maintaining low salt concentrations (150mM) to preserve electrostatic interactions. Indeed, it was not our aim to resolve stringent core molecular complexes, which is the scope of many large-scale protein interaction screens in the literature that constitute a significant fraction of the InWeb dataset. These studies, for the most part, use yeast two-hybrid assays and pull-downs of tagged proteins in overexpression systems. Rather, our aims were to map the brain cell-type-specific cellular networks anchored in index proteins linked to schizophrenia, use these inclusive datasets to model pathways and networks in the disease, and integrate with genetic datasets that are otherwise difficult to interpret mechanistically.

3. To further illustrate the importance of cell-type-specificity in our approach, we compared the index protein interactors identified in our study vs. those already published in InWeb in more detail (see table below). In general, we found a lower percentage of overlap with InWeb for index proteins that have more neuron-specific functions and expression (e.g., 1.3% for SYNGAP1), and a higher percentage of overlap for index proteins with more ubiquitous expression across tissues and cell types (e.g., 45.2% for CUL3). This indicates that most of the 'novel' interactions in our data are driven by neuronal genes that are underrepresented in InWeb, which is generally enriched for genes that are more amenable to biochemical assays in proliferative, non-neuronal cell models (e.g., HEK cells). These observations are in agreement with results from our previously published Genoppi paper (*18*), in which we applied a similar IP-MS protocol to identify interactors for four unrelated proteins (BCL2, TDP-43, MDM2, and PTEN) across four neuronal or cancer cell lines: we found that only ~17% of the interactions were reported in InWeb and that they clustered according to the

origin of the cell line used in the IPs, with a clear set of cell-type-specific interactors both in neurons and in various cancer cell lines.

| **Gene** | **# of interactors in this study** | **# of interactors in InWeb** | **# in overlap** | **overlap/this study %** | **overlap/InWeb %** |
|---|---|---|---|---|---|
| CACNA1C | 636 | 65 | 15 | 2.36 | 23.08 |
| CUL3 | 62 | 1266 | 28 | 45.16 | 2.21 |
| HCN1 | 673 | 8 | 5 | 0.74 | 62.50 |
| RIMS1 | 245 | 102 | 16 | 6.53 | 15.69 |
| SYNGAP1 | 375 | 36 | 5 | 1.33 | 13.89 |
| TCF4 | 26 | 270 | 3 | 11.54 | 1.11 |

Finally, we also assessed whether the computational approach (Genoppi/limma) we used to analyze the IP-MS data contributes significantly to the large portion of novel interactions we identified. In the Genoppi paper, we delved into this issue by analyzing the IP-MS data using both Genoppi and an alternative established method, SAINTexpress (*70*), and found that on average ~85% of the interactors identified by Genoppi were also identified by SAINTexpress, indicating strong agreement between the two methods. This shows that our specific computational method for analyzing the data is not driving the number of new interactions reported in this study.

In conclusion, most newly reported interactions in the current study likely result from: i) an experimental design that aims at capturing endogenous protein interactions, ii) a deliberately inclusive IP protocol to identify neuronal networks and pathways rather than exclusively stringent core molecular complexes [which is the data type typically reported in InWeb], and iii) an experimental system to capture brain cell-type-specific protein interactions that have not been studied in previous literature.

### Figures S1-S11

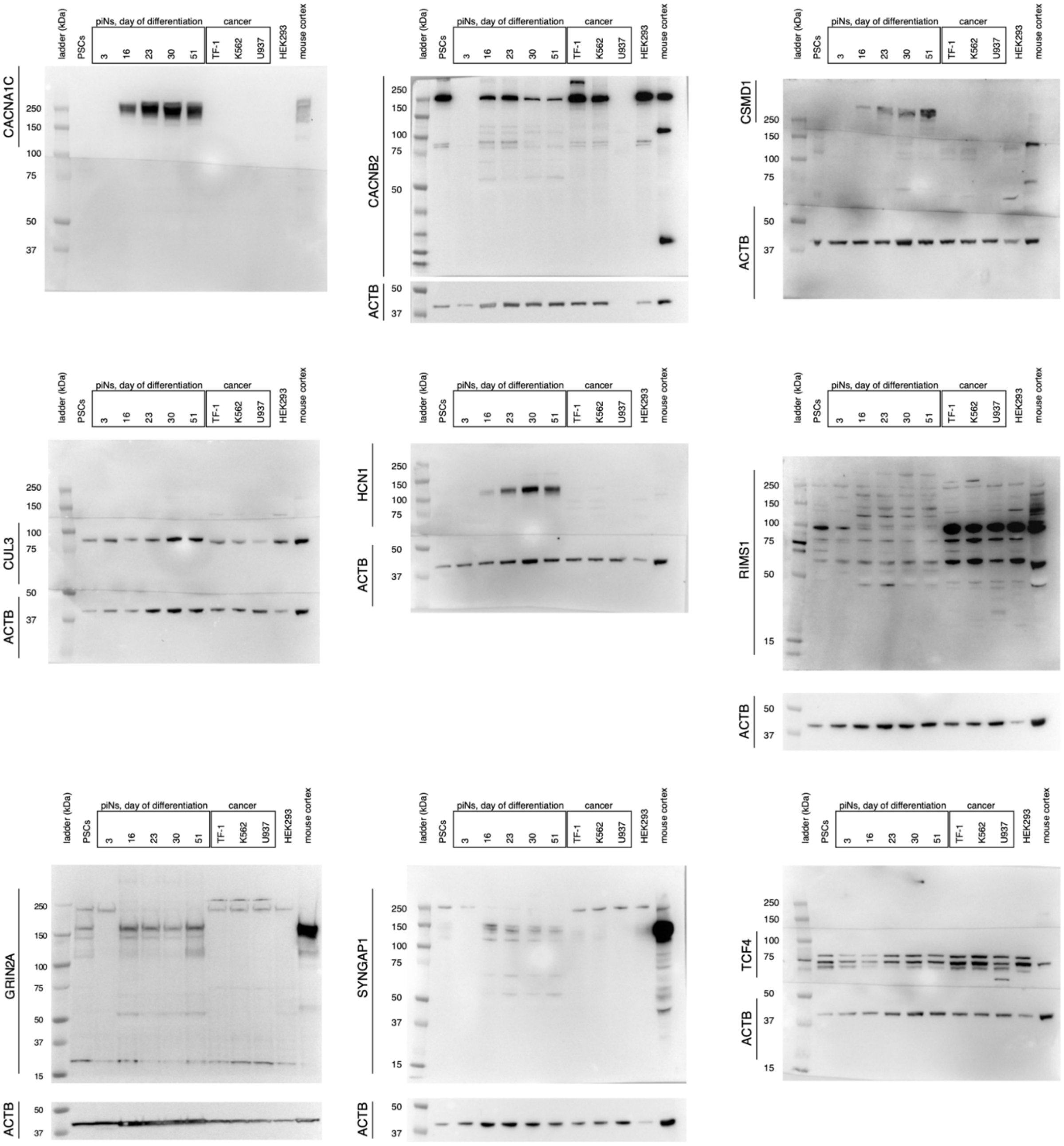


**Fig. S1. Western blot analysis of index proteins [corresponds to Fig. 1D].**

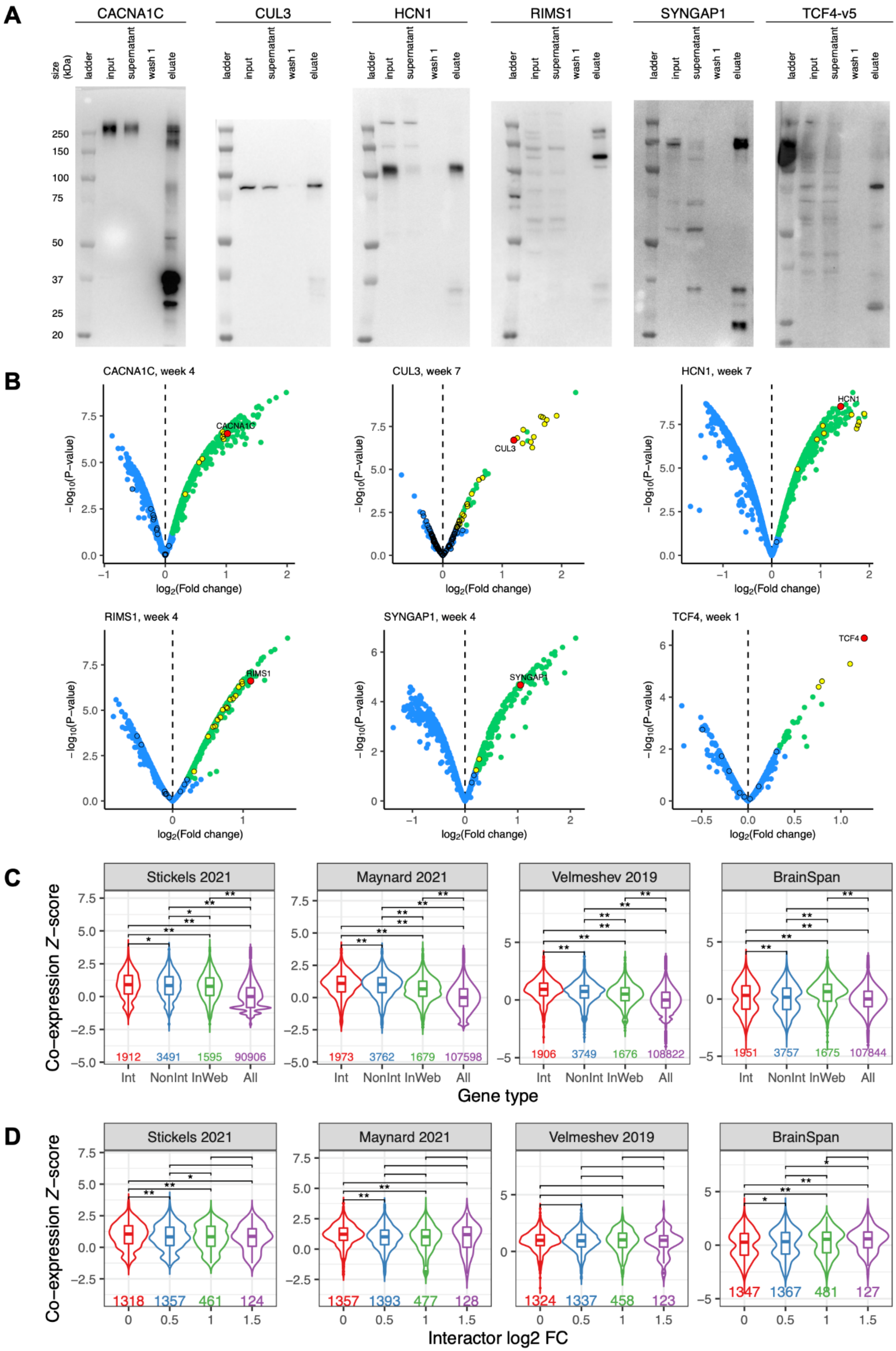
A
CACNA1C
CUL3
HCN1
RIMS1
SYNGAP1
TCF4-v5
size (kDa)
ladder
input
supernatant
wash 1
eluate
250
150
100
75
50
37
25
20
B
CACNA1C, week 4
CUL3, week 7
HCN1, week 7
RIMS1, week 4
SYNGAP1, week 4
TCF4, week 1
CACNA1C
CUL3
HCN1
RIMS1
SYNGAP1
TCF4
$-\log_{10}$(P−value)
$\log_2$(Fold change)
C
Stickels 2021
Maynard 2021
Velmeshev 2019
BrainSpan
Co−expression Z−score
1912
3491
1595
90906
1973
3762
1679
107598
1906
3749
1676
108822
1951
3757
1675
107844
Int
NonInt
InWeb
All
Gene type
D
Stickels 2021
Maynard 2021
Velmeshev 2019
BrainSpan
Co−expression Z−score
1318
1357
461
124
1357
1393
477
128
1324
1337
458
123
1347
1367
481
127
Interactor log2 FC

**Fig. S2. IP-MS experiments of six index proteins. (A)** Representative western blot analysis on IPs of the index proteins. Input, 10% of the protein lysate utilized for the IP; supernatant, 10% of unbound lysate; wash 1, 10% of eluate following the first wash; eluate, immunoprecipitate. **(B)** Representative volcano plots of IP-MS experiments of the index proteins. The index protein is shown in red, significant interactors [$\log_2$ FC > 0 and FDR ≤ 0.1] in green, and other detected proteins in blue. Known InWeb interactors are indicated by black border circles, with the subset that are significant in the IP highlighted in yellow. **(C)** Pairwise co-expression Z-scores between index genes and their interactors [Int], non-interactors [NonInt], known InWeb interactors [InWeb], and all protein-coding genes [All]. Scores were calculated from spatial transcriptomic datasets in mouse neocortex [Stickels 2021] and human dorsolateral prefrontal cortex [Maynard 2021], single-cell RNA-seq dataset in human cortex [Velmeshev 2019], and the BrainSpan RNA-seq dataset. Single or double asterisks indicate nominally [$P < 0.05$] or Bonferroni [$P < 0.05/6$, adjusting for 6 pairwise comparisons] significant difference in score distribution, respectively, as calculated by two-tailed Wilcoxon rank-sum tests. Number of gene pairs plotted for each gene type is indicated towards the bottom of the plot. **(D)** Pairwise co-expression Z-scores between index genes and their interactors identified at different $\log_2$ FC thresholds, following same annotations as (C).

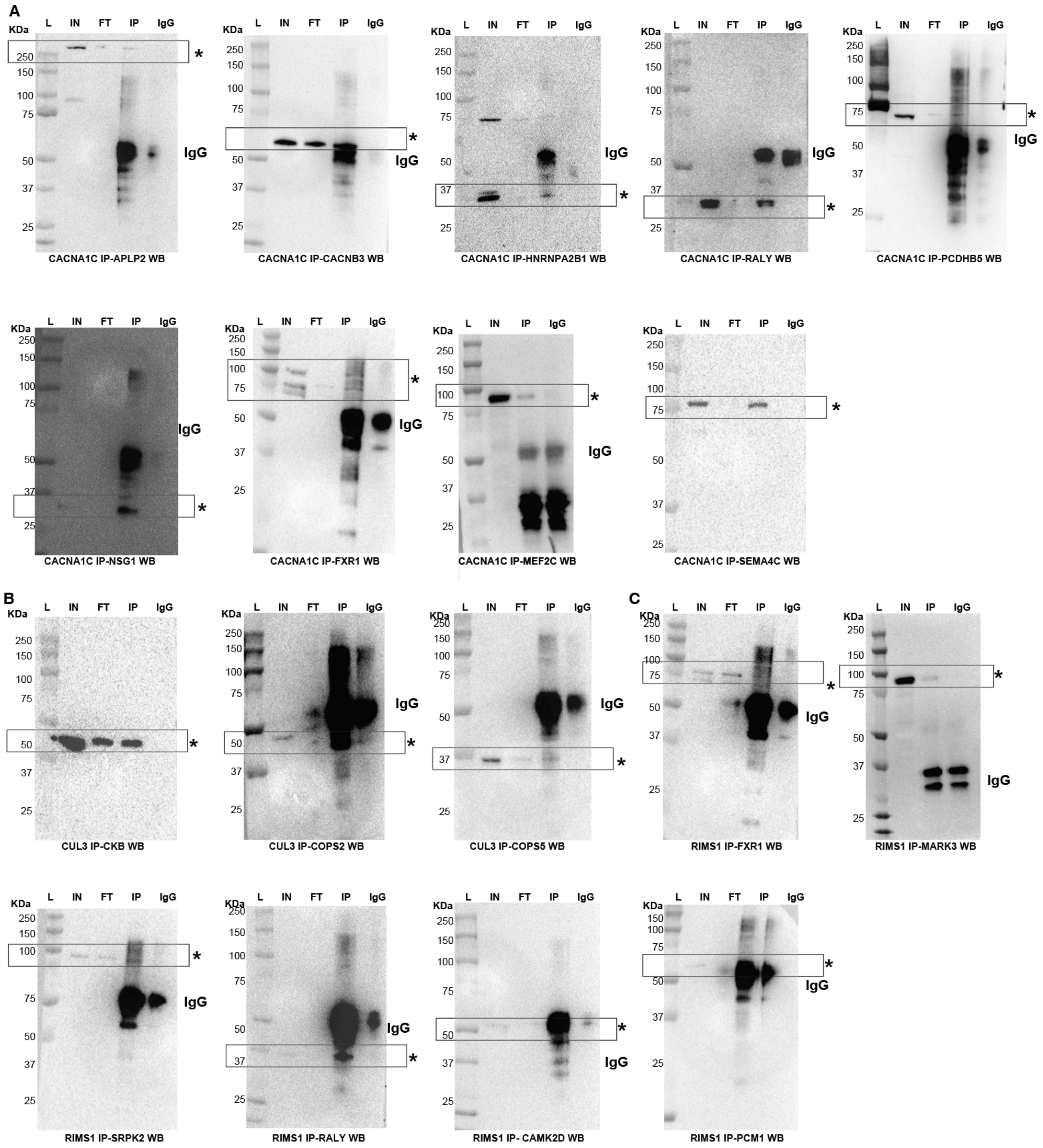


**Fig. S3. Western blot analysis on IPs of index proteins to detect the presence of selected interactors.** **(A)** CACNA1C immunoprecipitations; **(B)** CUL3 immunoprecipitations; **(C)** RIMS1 immunoprecipitations; **(D)** SYNGAP1 immunoprecipitations. Molecular weights are in KDa. L=Ladder, IN=Input, FT=Flow-through, IP=Immunoprecipitation, IgG=IgG control. An asterisk marks the expected band(s) of each named protein. Boxes highlight the molecular weight where expected signal should be compared across lanes. All validation experiments were performed at week 4 of neuronal differentiation.

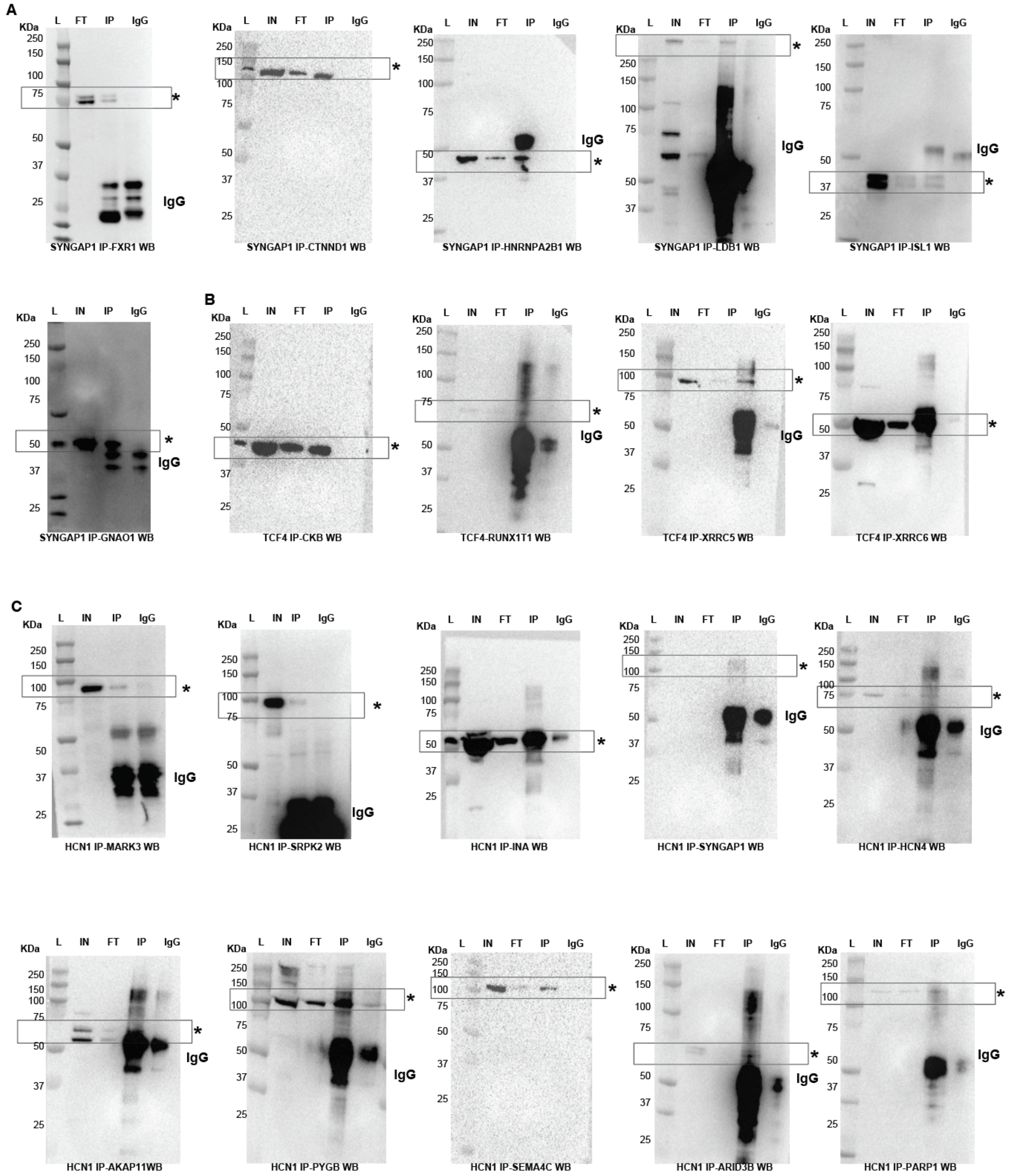


**Fig. S4. Western blot analysis on IPs of index proteins to detect the presence of selected interactors.** **(A)** SYNGAP1 immunoprecipitations; **(B)** TCF4 immunoprecipitations; **(C)** HCN1 immunoprecipitations. Molecular weights are in KDa. L=Ladder, IN=Input, FT=Flow-through, IP=Immunoprecipitation, IgG=IgG control. An asterisk marks the expected band(s) of each named protein. Boxes highlight the molecular weight where expected signal should be compared across lanes. All validation experiments were performed at week 4 of neuronal differentiation.

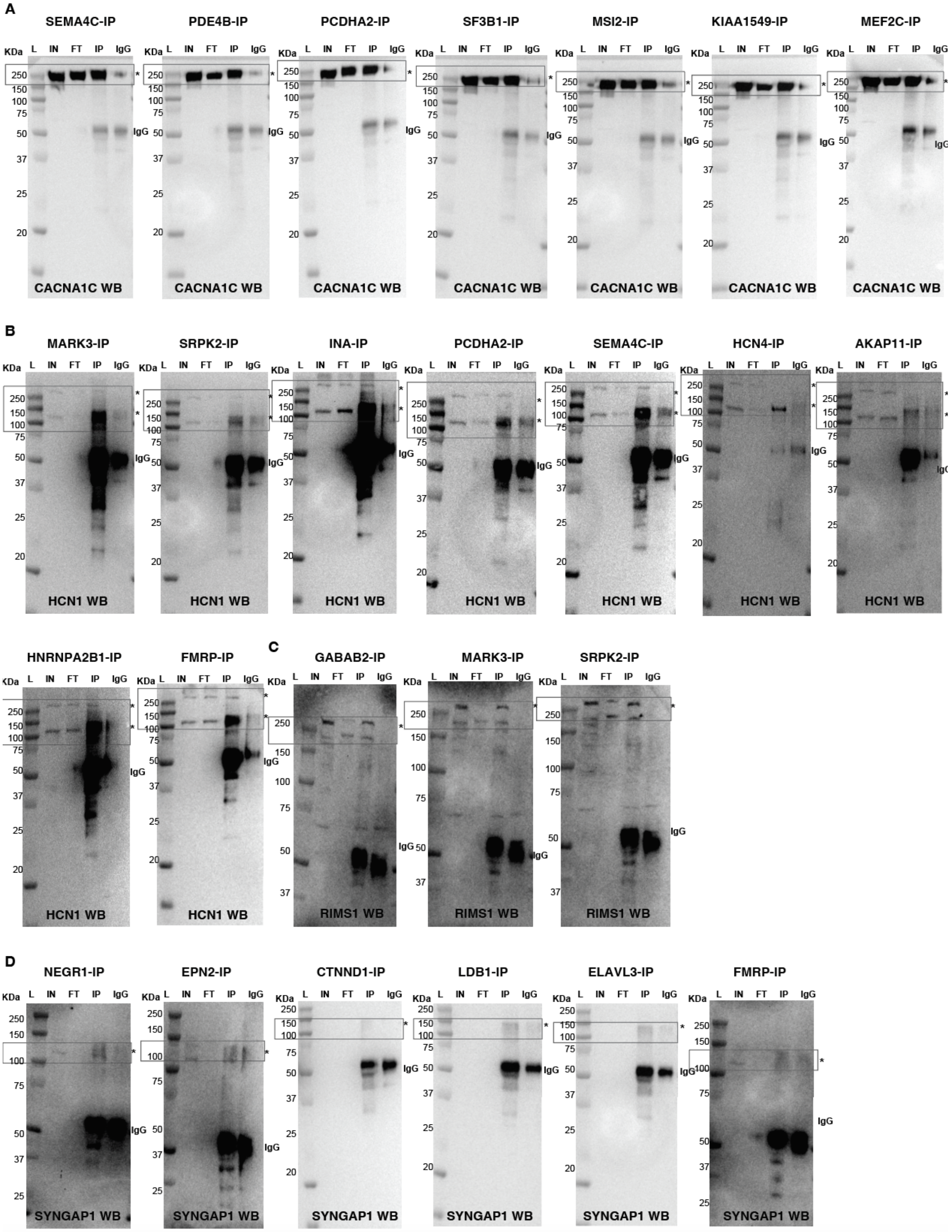
A
SEMA4C-IP
PDE4B-IP
PCDHA2-IP
SF3B1-IP
MSI2-IP
KIAA1549-IP
MEF2C-IP
KDa L IN FT IP IgG
250
150
100
75
50
37
25
20
IgG
CACNA1C WB
B
MARK3-IP
SRPK2-IP
INA-IP
PCDHA2-IP
SEMA4C-IP
HCN4-IP
AKAP11-IP
HNRNPA2B1-IP
FMRP-IP
HCN1 WB
C
GABAB2-IP
MARK3-IP
SRPK2-IP
RIMS1 WB
D
NEGR1-IP
EPN2-IP
CTNND1-IP
LDB1-IP
ELAVL3-IP
FMRP-IP
SYNGAP1 WB

**Fig. S5. Western blot analysis on reverse IPs of selected interactors to detect the presence of index proteins. (A)** CACNA1C western blots; **(B)** HCN1 western blots. HCN1 runs as a monomer or dimer, at ~190KDa and ~380KDa. Depending on the reverse IP, either one or both bands are present in the IP lane; **(C)** RIMS1 western blots; **(D)** SYNGAP1 western blots. Due to low levels of expression of neuronal TDP-43, we could only detect a dim signal in the input for SYNGAP1 prior to enrichment by IP. Molecular weights are in KDa. L=Ladder, IN=Input [10% of the protein lysate utilized for the IP]; FT=supernatant [10% of unbound lysate]; IP=immunoprecipitate; IgG=IgG control. An asterisk marks the expected band(s) of each named protein. Boxes highlight the molecular weight where expected signal should be compared across lanes. The name of the immunoprecipitated protein is on top of each blot. All validation experiments were performed at week 4 of neuronal differentiation.

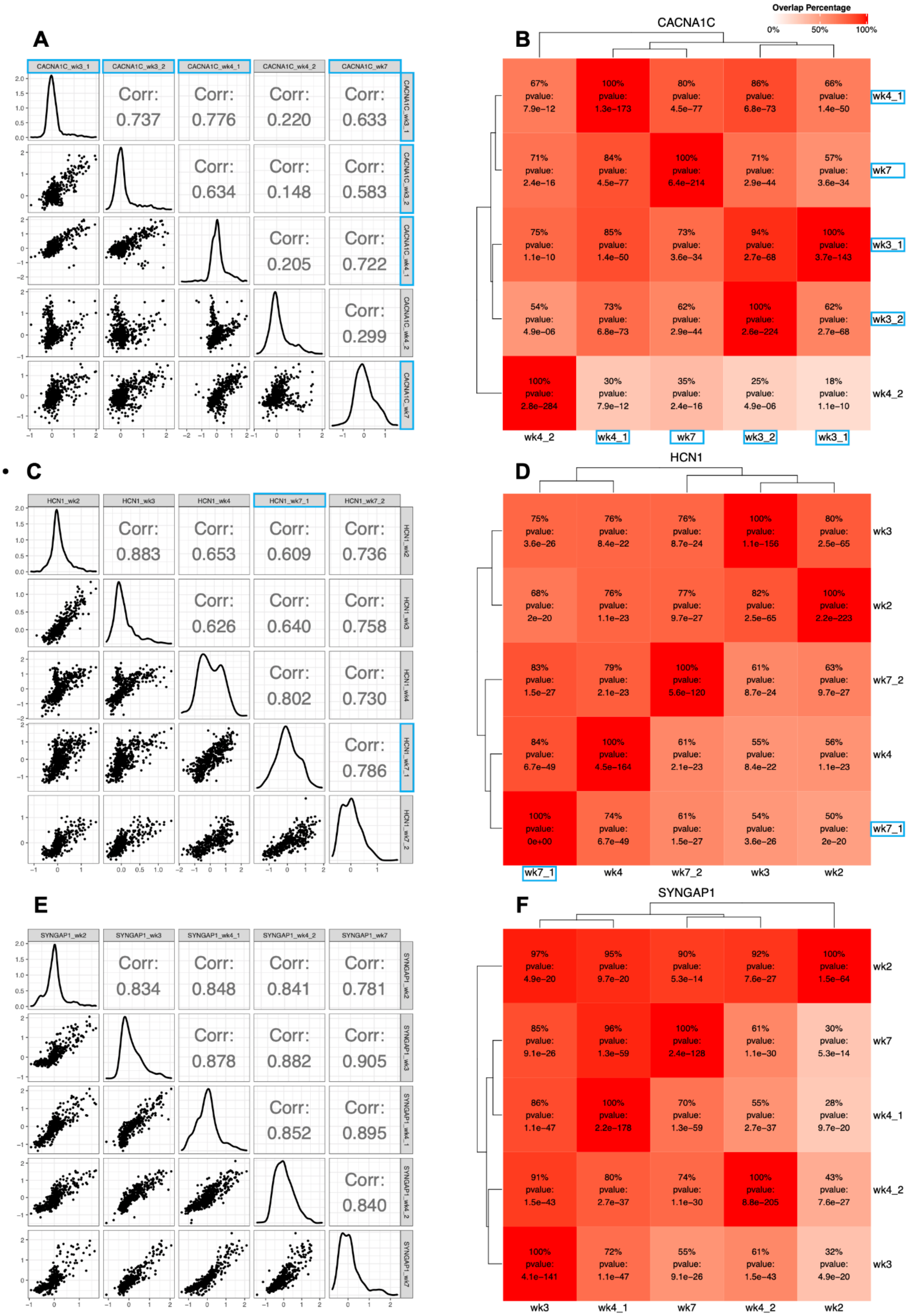
A
CACNA1C_wk3_1
CACNA1C_wk3_2
CACNA1C_wk4_1
CACNA1C_wk4_2
CACNA1C_wk7
Corr: 0.737
Corr: 0.776
Corr: 0.220
Corr: 0.633
Corr: 0.634
Corr: 0.148
Corr: 0.583
Corr: 0.205
Corr: 0.722
Corr: 0.299
B
CACNA1C
Overlap Percentage
0%
50%
100%
67% pvalue: 7.9e−12
100% pvalue: 1.3e−173
80% pvalue: 4.5e−77
86% pvalue: 6.8e−73
66% pvalue: 1.4e−50
wk4_1
71% pvalue: 2.4e−16
84% pvalue: 4.5e−77
100% pvalue: 6.4e−214
71% pvalue: 2.9e−44
57% pvalue: 3.6e−34
wk7
75% pvalue: 1.1e−10
85% pvalue: 1.4e−50
73% pvalue: 3.6e−34
94% pvalue: 2.7e−68
100% pvalue: 3.7e−143
wk3_1
54% pvalue: 4.9e−06
73% pvalue: 6.8e−73
62% pvalue: 2.9e−44
100% pvalue: 2.6e−224
62% pvalue: 2.7e−68
wk3_2
100% pvalue: 2.8e−284
30% pvalue: 7.9e−12
35% pvalue: 2.4e−16
25% pvalue: 4.9e−06
18% pvalue: 1.1e−10
wk4_2
wk4_2
wk4_1
wk7
wk3_2
wk3_1
C
HCN1_wk2
HCN1_wk3
HCN1_wk4
HCN1_wk7_1
HCN1_wk7_2
Corr: 0.883
Corr: 0.653
Corr: 0.609
Corr: 0.736
Corr: 0.626
Corr: 0.640
Corr: 0.758
Corr: 0.802
Corr: 0.730
Corr: 0.786
D
HCN1
75% pvalue: 3.6e−26
76% pvalue: 8.4e−22
76% pvalue: 8.7e−24
100% pvalue: 1.1e−156
80% pvalue: 2.5e−65
wk3
68% pvalue: 2e−20
76% pvalue: 1.1e−23
77% pvalue: 9.7e−27
82% pvalue: 2.5e−65
100% pvalue: 2.2e−223
wk2
83% pvalue: 1.5e−27
79% pvalue: 2.1e−23
100% pvalue: 5.6e−120
61% pvalue: 8.7e−24
63% pvalue: 9.7e−27
wk7_2
84% pvalue: 6.7e−49
100% pvalue: 4.5e−164
61% pvalue: 2.1e−23
55% pvalue: 8.4e−22
56% pvalue: 1.1e−23
wk4
100% pvalue: 0e+00
74% pvalue: 6.7e−49
61% pvalue: 1.5e−27
54% pvalue: 3.6e−26
50% pvalue: 2e−20
wk7_1
wk7_1
wk4
wk7_2
wk3
wk2
E
SYNGAP1_wk2
SYNGAP1_wk3
SYNGAP1_wk4_1
SYNGAP1_wk4_2
SYNGAP1_wk7
Corr: 0.834
Corr: 0.848
Corr: 0.841
Corr: 0.781
Corr: 0.878
Corr: 0.882
Corr: 0.905
Corr: 0.852
Corr: 0.895
Corr: 0.840
F
SYNGAP1
97% pvalue: 4.9e−20
95% pvalue: 9.7e−20
90% pvalue: 5.3e−14
92% pvalue: 7.6e−27
100% pvalue: 1.5e−64
wk2
85% pvalue: 9.1e−26
96% pvalue: 1.3e−59
100% pvalue: 2.4e−128
61% pvalue: 1.1e−30
30% pvalue: 5.3e−14
wk7
86% pvalue: 1.1e−47
100% pvalue: 2.2e−178
70% pvalue: 1.3e−59
55% pvalue: 2.7e−37
28% pvalue: 9.7e−20
wk4_1
91% pvalue: 1.5e−43
80% pvalue: 2.7e−37
74% pvalue: 1.1e−30
100% pvalue: 8.8e−205
43% pvalue: 7.6e−27
wk4_2
100% pvalue: 4.1e−141
72% pvalue: 1.1e−47
55% pvalue: 9.1e−26
61% pvalue: 1.5e−43
32% pvalue: 4.9e−20
wk3
wk3
wk4_1
wk7
wk4_2
wk2

**Fig. S6. Comparison of CACNA1C, HCN1, and SYNGAP1 IPs across neuron differentiation time points and cell lines. (A)** Scatter plots and Pearson's correlations comparing $\log_2$ FC values of detected proteins across CACNA1C IPs. **(B)** Clustered heat map showing the overlap of significant interactors between each pair of CACNA1C IPs. Overlap percentages were calculated using the row's IP experiment as the denominator; the corresponding overlap enrichment P-values were calculated using a one-tailed hypergeometric test. Dendrograms were generated from hierarchical clustering of the overlap percentages as implemented in the ComplexHeatmap R package (*71*) (v2.2.0). **(C)-(D)** The analogous panels for HCN1 IPs. **(E)-(F)** The analogous panels for SYNGAP1 IPs. Neuron differentiation time point of each IP is indicated by the 'wk' abbreviation in the IP name label [e.g., 'wk3' = week 3]. IPs performed using iNs derived from the WA01 [H1] cell line are boxed in blue; all other IPs were performed using iNs derived from the iPS3 cell line.

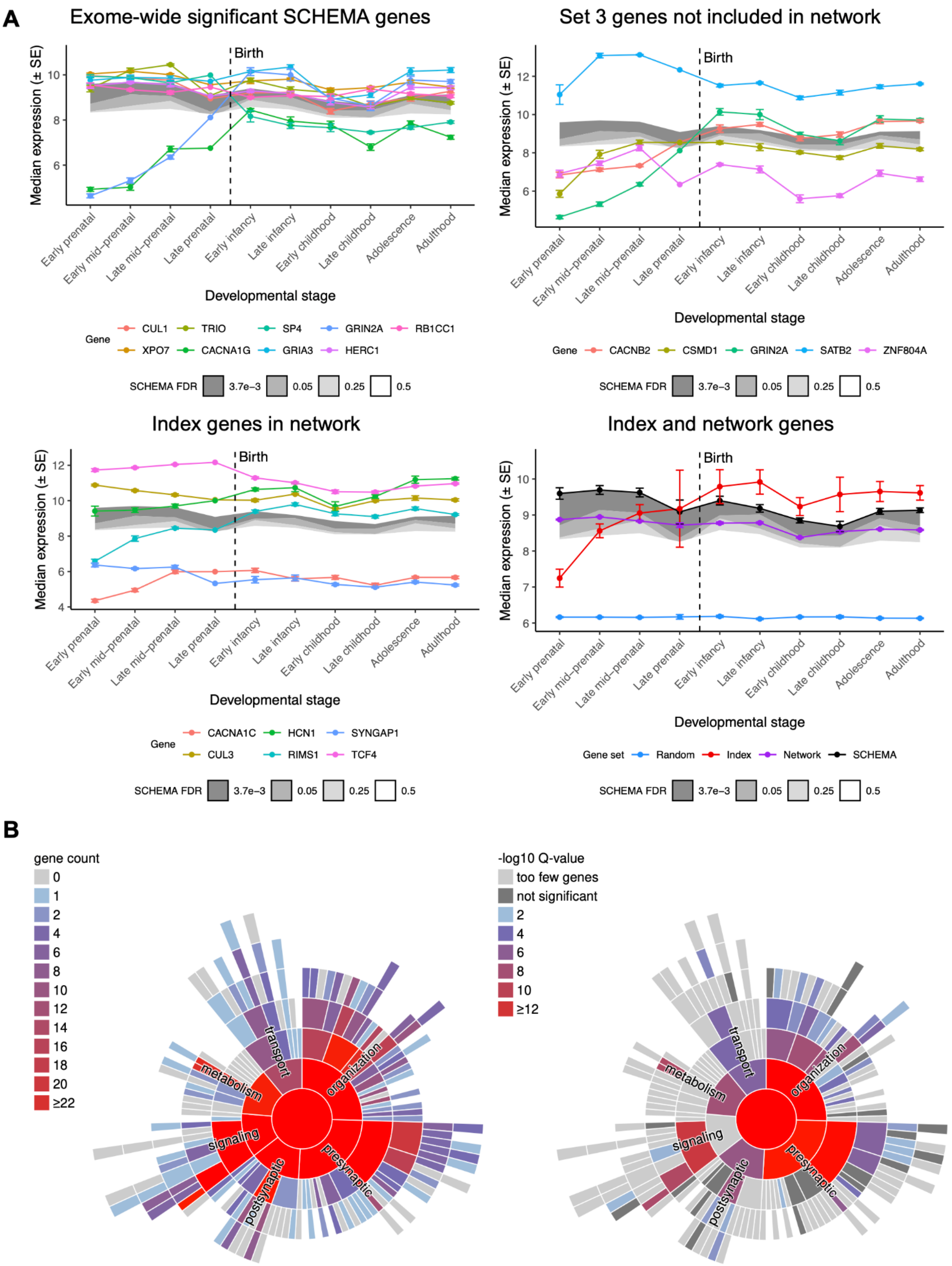
A
Exome-wide significant SCHEMA genes
Set 3 genes not included in network
Index genes in network
Index and network genes
Birth
Median expression (± SE)
Developmental stage
Early prenatal
Early mid−prenatal
Late mid−prenatal
Late prenatal
Early infancy
Late infancy
Early childhood
Late childhood
Adolescence
Adulthood
Gene
CUL1
TRIO
SP4
GRIN2A
RB1CC1
XPO7
CACNA1G
GRIA3
HERC1
CACNB2
CSMD1
SATB2
ZNF804A
CACNA1C
HCN1
SYNGAP1
CUL3
RIMS1
TCF4
Gene set
Random
Index
Network
SCHEMA
SCHEMA FDR
3.7e−3
0.05
0.25
0.5
B
gene count
≥22
-log10 Q-value
too few genes
not significant
≥12
transport
organization
metabolism
signaling
presynaptic
postsynaptic

**Fig. S7. BrainSpan expression and SynGO enrichment of the all combined network. (A)** Frontal cortex RNA expression of individual exome-wide significant SCHEMA genes [upper left], Set 3 genes not included in our interaction network [upper right], index genes included in the network [lower left], and all network genes [lower right] across ten developmental stages (see **Text S4** for discussion). Median expression and standard error [SE] were derived from the BrainSpan exon microarray dataset. 'Random' indicates genes randomly sampled from the BrainSpan dataset; 'Index' indicates the six index genes; 'Network' indicates genes encoding the interactors in the network; 'SCHEMA' indicates exome-wide significant genes from SCHEMA. Shaded regions indicate median expression of genes with FDR < 3.7e-3 [exome-wide significance], 0.05, 0.25, or 0.5 in SCHEMA with darker grey indicating greater significance. **(B)** SynGO analysis of genes in the interaction network. The sunburst plots show the number [left] and the enrichment [right] of genes distributed over a range of biological processes in the synapse.

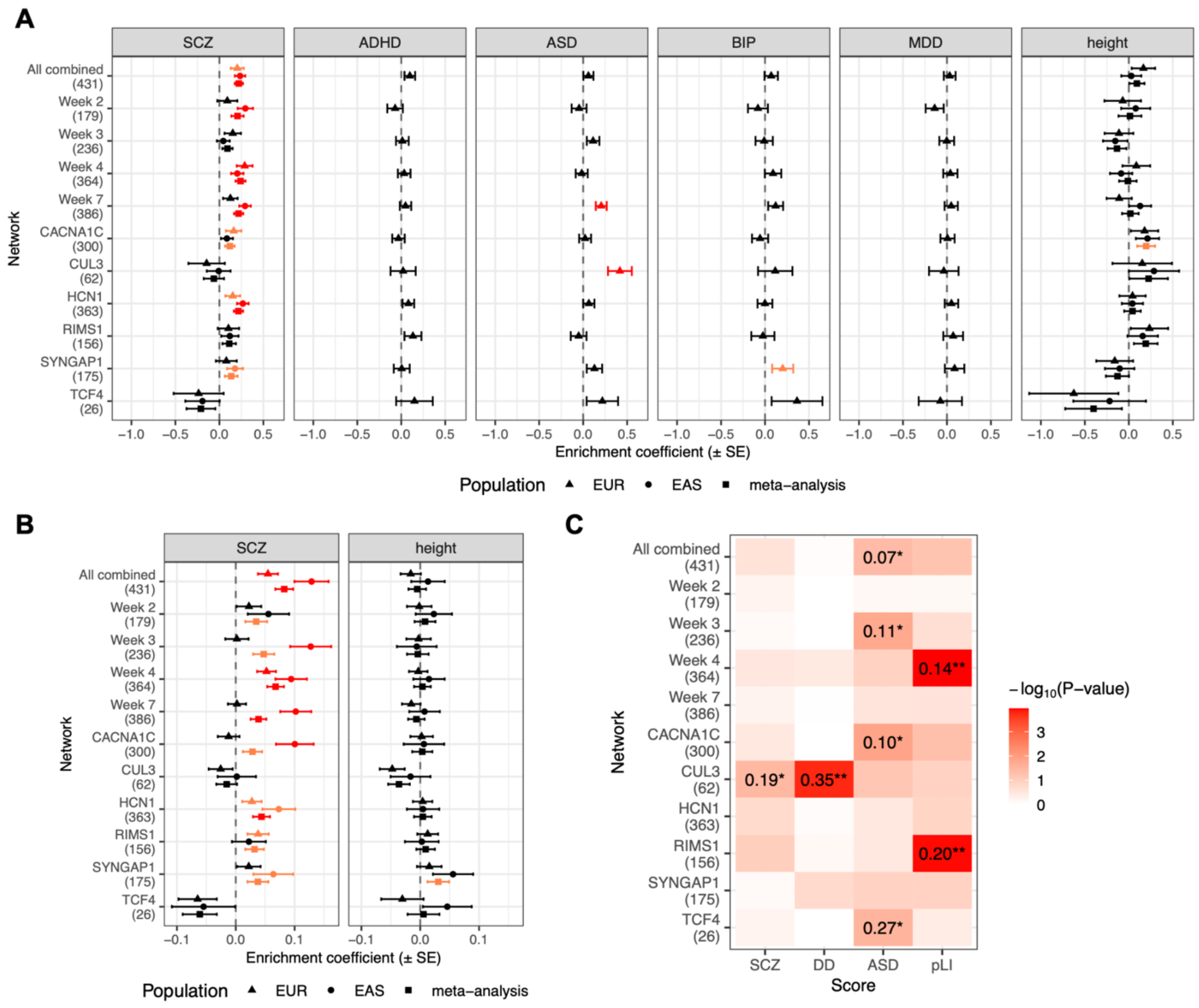


**Fig. S8. Genetic risk enrichment in the index protein interactomes.** Networks tested are the combined network of all IPs [All combined], the combined networks at each time point [Week 2 to Week 7], the combined networks for CACNA1C, HCN1, RIMS1, and SYNGAP1, and the individual IP networks for CUL3 and TCF4; the number of genes in each network is shown in parentheses on the y-axes. **(A)-(B)** Common variant enrichment of schizophrenia [SCZ], attention deficit hyperactivity disorder [ADHD], autism spectrum disorders [ASD], bipolar disorder [BIP], major depressive disorder [MDD], or height calculated using MAGMA **(A)** or the GRS method **(B)** and GWAS data of European [EUR] or East Asian [EAS] ancestry. Nominal [$P < 0.05$] or Bonferroni [$P < 0.05/22$, adjusting for 11 networks and two ancestries] significance is highlighted in orange or red, respectively. **(C)** Enrichment of rare variant-based SCZ, developmental disorders [DD], or ASD association statistics or gnomAD pLI scores calculated using one-tailed KS tests. KS test statistics reaching nominal or Bonferroni [$P <$

0.05/11, adjusting for 11 networks] significance are shown in the heat map followed by single or double asterisks, respectively.

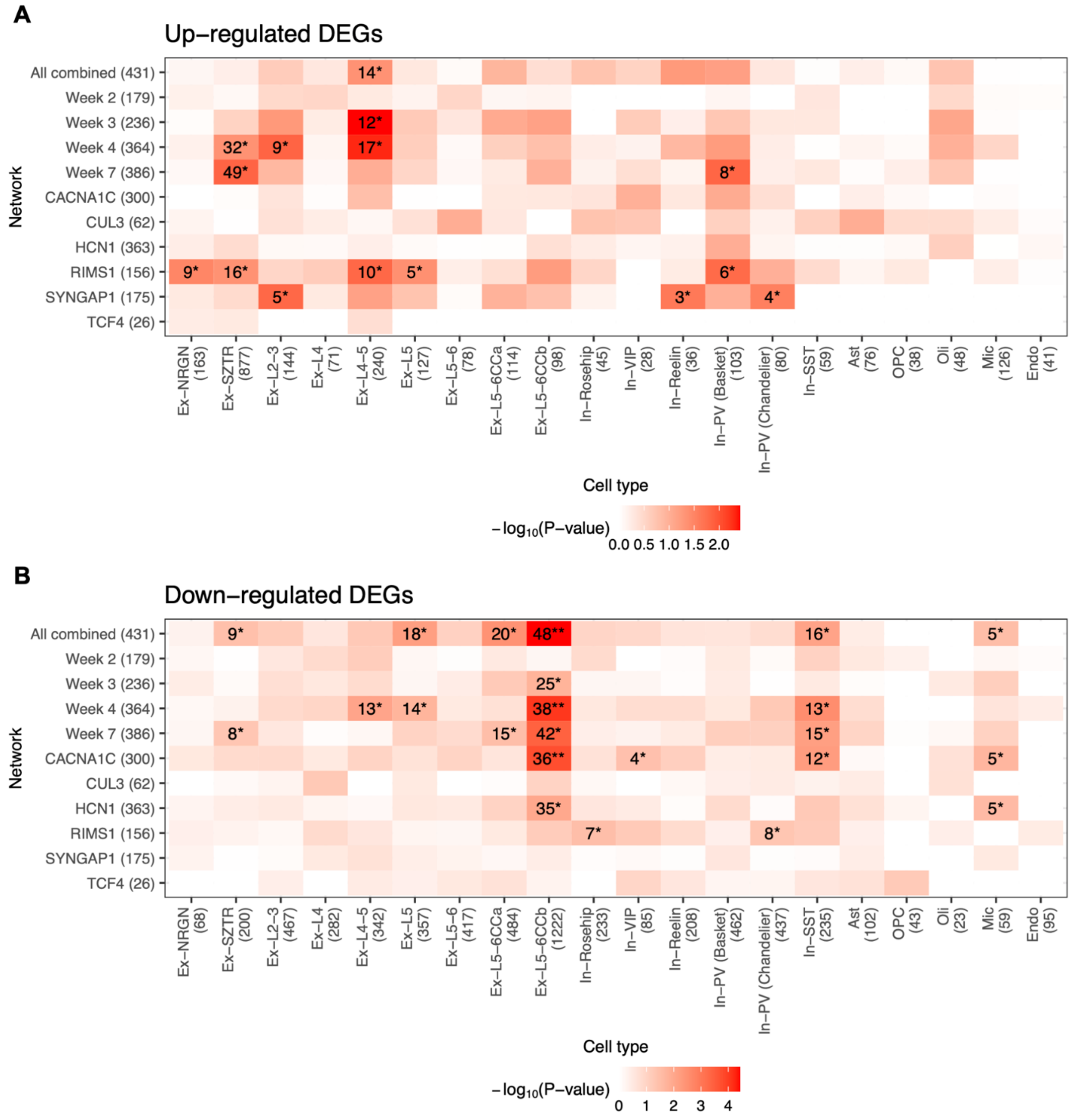


**Fig. S9. Enrichment of cell-type-specific differentially expressed genes [DEGs] in the prefrontal cortex of schizophrenia patients in the index protein interactomes.** Networks tested are the combined network of all IPs [All combined], the combined networks at each time point [Week 2 to Week 7], the combined networks for CACNA1C, HCN1, RIMS1, and SYNGAP1, and the individual IP networks for CUL3 and TCF4; the number of genes in each network is shown in parentheses on the y-axes. Enrichment of up-regulated **(A)** and down-regulated **(B)** DEGs are shown separately; the number of DEGs in each cell type is shown in parentheses on the x-axes. P-values were calculated using one-tailed hypergeometric tests. Gene counts in overlaps reaching nominal or Bonferroni [$P < 0.05/220$, adjusting for 11 networks and 20 cell types] significance are shown in the heat maps followed by single or double asterisks, respectively.

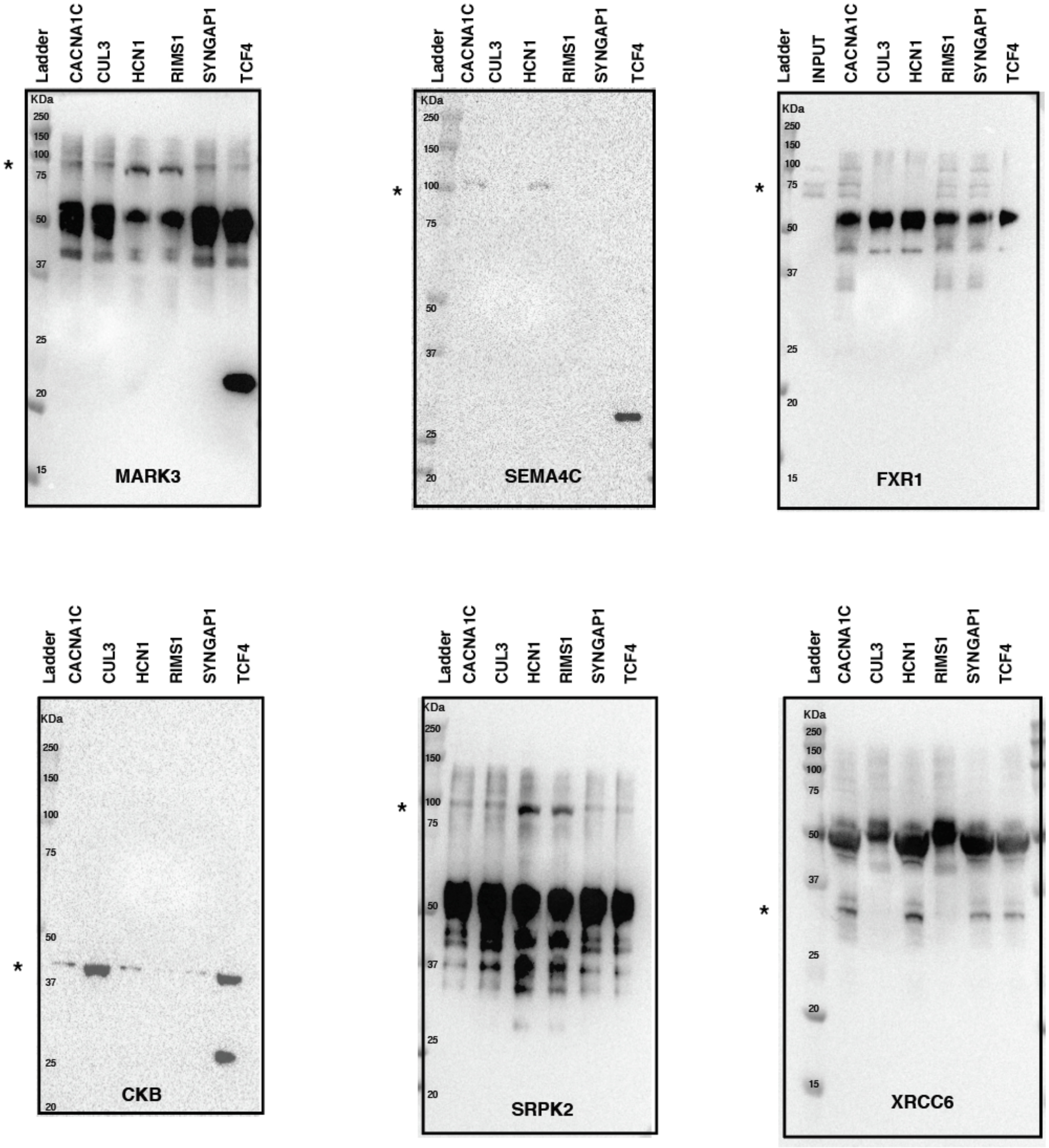


**Fig. S10. Western blot analysis on IPs of index proteins (panels) to detect the presence of selected interactors found in GWAS loci [corresponds to Fig. 4D].** Immunoprecipitations of all index proteins (named at the top of each gel) were performed at week 4 of neuronal differentiation. An asterisk indicates the expected molecular weight of each blotted interactor (named at the bottom of each gel).

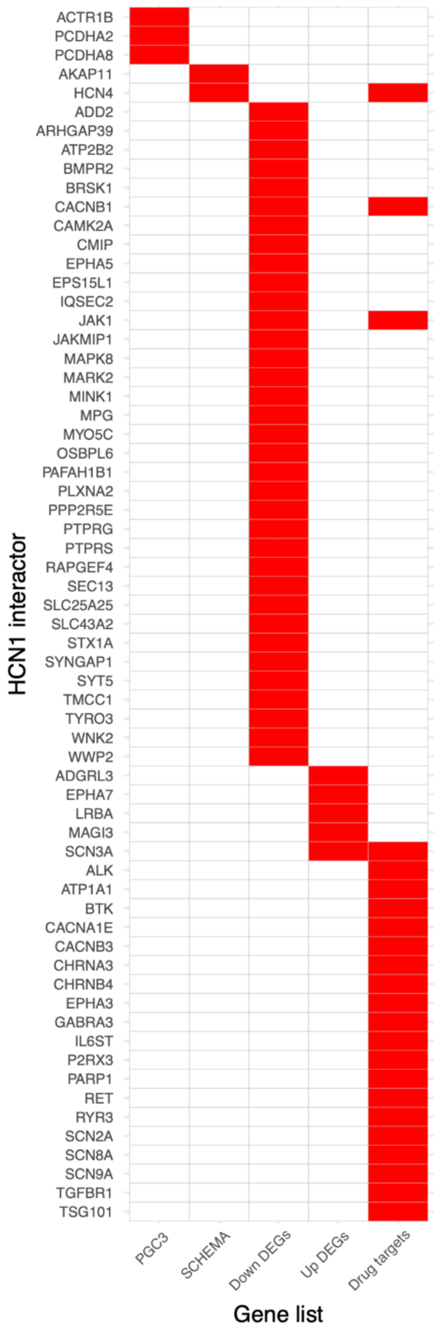


**Fig. S11. Overlap [red] between the combined HCN1 network ['stringent interactors' in Data S9] vs. various gene lists.** 'PGC3' indicates genes prioritized using FINEMAP or SMR approach in the PGC schizophrenia GWAS [phase 3] paper; 'SCHEMA' indicates genes with FDR < 0.05 from SCHEMA; 'Down DEGs' and 'Up DEGs' indicate down- or up-regulated DEGs, respectively, in the 'Ex-L5-6CCb' cell type in schizophrenia patients; 'Drug targets' indicates genes targeted by existing drugs.

**Captions for Data S1-S14**

**Data S1. Details of index gene selection procedure.**

**Data S2. Enrichment of pLI scores in schizophrenia gene sets calculated using KS test.**

**Data S3. BrainSpan expression profiles of schizophrenia gene sets.**

**Data S4. Antibodies tested and used in this study.**

**Data S5. Overview of the 19 IP-MS experiments that passed QC.**

**Data S6. Genoppi analysis results for the 19 IP-MS experiments that passed QC.**

**Data S7. Summary of interactions tested in forward or reverse IP-WB validation experiments.**

**Data S8. Overlap enrichment of significant interactors across IPs of the same index protein.**

**Data S9. Interactor and non-interactor genes in the 19 individual IP and the 9 combined datasets.**

**Data S10. SynGO gene set analysis for the all combined network.**

**Data S11. Genetic risk enrichment in the interaction networks.**

**Data S12. Enrichment of cell-type-specific differentially expressed genes (DEGs) in the prefrontal cortex of schizophrenia patients in the interaction networks.**

**Data S13. Protein-coding genes in PGC schizophrenia GWAS (phase 3) loci prioritized by FINEMAP, SMR, or our interaction data.**

**Data S14. Drug targets in the HCN1 network.**
